# Changes in the trajectory of Long Covid symptoms following COVID-19 vaccination: community-based cohort study

**DOI:** 10.1101/2021.12.09.21267516

**Authors:** Daniel Ayoubkhani, Charlotte Bermingham, Koen B. Pouwels, Myer Glickman, Vahé Nafilyan, Francesco Zaccardi, Kamlesh Khunti, Nisreen A. Alwan, A. Sarah Walker

## Abstract

**Objective:** To estimate associations between COVID-19 vaccination and Long Covid symptoms in adults who were infected with SARS-CoV-2 prior to vaccination.

**Design:** Observational cohort study using individual-level interrupted time series analysis.

**Setting:** Random sample from the community population of the UK.

**Participants:** 28,356 COVID-19 Infection Survey participants (mean age 46 years, 56% female, 89% white) aged 18 to 69 years who received at least their first vaccination after test-confirmed infection.

**Main outcome measures:** Presence of long Covid symptoms at least 12 weeks after infection over the follow-up period 3 February to 5 September 2021.

**Results:** Median follow-up was 141 days from first vaccination (among all participants) and 67 days from second vaccination (84% of participants). First vaccination was associated with an initial 12.8% decrease (95% confidence interval: −18.6% to −6.6%) in the odds of Long Covid, but increasing by 0.3% (−0.6% to +1.2%) per week after the first dose. Second vaccination was associated with an 8.8% decrease (−14.1% to −3.1%) in the odds of Long Covid, with the odds subsequently decreasing by 0.8% (−1.2% to −0.4%) per week. There was no statistical evidence of heterogeneity in associations between vaccination and Long Covid by socio-demographic characteristics, health status, whether hospitalised with acute COVID-19, vaccine type (adenovirus vector or mRNA), or duration from infection to vaccination.

**Conclusions:** The likelihood of Long Covid symptoms reduced after COVID-19 vaccination, and the improvement was sustained over the follow-up period after the second dose. Vaccination may contribute to a reduction in the population health burden of Long Covid, though longer follow-up time is needed.

**Summary box:** What is already known on this topic

- COVID-19 vaccines are effective at reducing rates of SARS-CoV-2 infection, transmission, hospitalisation, and death
- The incidence of Long Covid may be reduced if infected after vaccination, but the relationship between vaccination and pre-existing long COVID symptoms is unclear, as published studies are generally small and with self-selected participants

What this study adds

- The likelihood of Long Covid symptoms reduced after COVID-19 vaccination, and the improvement was sustained over the follow-up period after the second dose
- There was no evidence of differences in this relationship by socio-demographic characteristics, health-related factors, vaccine type, or duration from infection to vaccination
- Although causality cannot be inferred from this observational evidence, vaccination may contribute to a reduction in the population health burden of Long Covid; further research is needed to understand the biological mechanisms that may ultimately contribute to the development of therapeutics for Long Covid

## Introduction

Symptoms may persist for months following SARS-CoV-2 infection, defined in UK clinical guidelines [1] as ongoing symptomatic COVID-19 (signs and symptoms from 4 to 12 weeks post-onset) or post-COVID-19 syndrome (more than 12 weeks post-onset). These symptoms are collectively and commonly referred to as Long Covid. Long Covid is characterised by a range of symptoms across organ systems, including fatigue, shortness of breath, and cognitive impairment [2], often with undulating periods of wellness followed by relapse [3–5]. By February 2021, nearly 6% of adults in England may have experienced prolonged symptoms following coronavirus infection since the pandemic began [6], and 1.2 million people in private households in the UK (1.9%) were estimated to be reporting Long Covid symptoms in October 2021, with symptoms having a detrimental impact on the day-to-day activities of two-thirds of these individuals [2].

Population-level immunisation against COVID-19 began in the UK on 8 December 2020, and both adenovirus vector and messenger ribonucleic acid (mRNA) vaccines have demonstrated safety and efficacy in trials [7–10] and real-world effectiveness at reducing rates of infection [11–12], transmission [13], hospitalisation [14] and death [14–15]. However, the impact of vaccination on Long Covid symptoms is uncertain. Preliminary research suggests that persistent symptoms are less common in breakthrough infections [16], but studies of people with pre-existing Long Covid have been small and with self-selected study participants [17–19].

We therefore used data from the Office for National Statistics (ONS) COVID-19 Infection Survey (CIS), a large, community-based population survey of randomly sampled UK households, to assess changes in the trajectory of Long Covid symptoms in adults infected with SARS-CoV-2 prior to receiving their first vaccination, and heterogeneity in trajectories by socio-demographics, health-related factors, vaccine type, and duration since infection.

## Methods

### Study data

Data were obtained from the CIS (ISRCTN21086382, www.ndm.ox.ac.uk/covid-19/covid-19-infection-survey/protocol-and-information-sheets) [20], a longitudinal survey of individuals aged 2 years or over in randomly sampled UK households (excluding communal establishments such as hospitals, care homes, halls of residence, and prisons), with ethical approval from the South Central Berkshire B Research Ethics Committee (20/SC/0195). After verbal agreement to participate, each selected household was visited by a study worker to provide written confirmed consent (from parents/carers for those aged 2 to 15 years; those aged 10 to 15 years also provided written assent). At the first visit, participants could consent for (optional) follow-up visits every week for the next month and then monthly for 12 months or longer. Supplementary Table 1 shows survey response rates.

All participants provided a nose and throat self-swab for polymerase chain reaction (PCR) testing at every follow-up visit. Individuals aged 16 years or older in a random subsample of households (initially 10% but expanded from April 2021), and those in households where another household member previously tested positive for SARS-CoV-2, were invited to provide monthly blood samples for S-antibody testing. Participants also reported whether they had tested positive for the virus or antibodies outside of the study (for example, through national testing programmes).

At every monthly visit since 3 February 2021, all CIS participants were asked whether they would describe themselves as currently experiencing Long Covid, defined as symptoms persisting for at least four weeks from confirmed or suspected coronavirus infection that could not be explained by another health condition. This definition uses self-classification of Long Covid, rather than a pre-specified symptoms list or clinical diagnosis, and thus reflects participants’ perception of whether their lived experience is consistent with what they understand of the condition. Participants who responded positively to the Long Covid question were further asked about the extent to which their day-to-day activities were limited as a result, and the presence of 21 individual symptoms as part of their experience of Long Covid (selected on the basis of being among the most commonly reported when the survey question was developed [4, 5, 21]; see Supplementary Table 2 for the full list).

For participants in England, vaccination information (number of doses, dates, manufacturer) was obtained from self-reported CIS responses and linked National Immunisation Management System (NIMS) records, with NIMS being prioritised where data conflicted. Concordance between self-reported and NIMS data was previously found to be high regarding vaccination type (98%) and date (95% within ±7 days) [11]. Administrative records were not available for participants in Wales, Scotland, and Northern Ireland, so vaccination data for these individuals were taken from the CIS alone.

### Study design

The analysis included CIS participants aged 18 to 69 years on 3 February 2021. Participants were included if they: responded to the survey question on Long Covid at least once up to 5 September 2021 (end of follow-up); received at least one COVID-19 vaccination before or during the follow-up period; and received a positive swab or blood test for SARS-CoV-2, either through the CIS or reported outside of the study, prior to vaccination. We excluded CIS participants remaining unvaccinated by 5 September 2021 because they were likely to differ from those who were vaccinated according to unmeasured characteristics (for example, personal considerations related to vaccine hesitancy).

### Infection date

Time of infection was the date of first positive swab or antibody test (ignoring blood tests after first vaccination), or the date when the participant first thought they had COVID-19 that was later confirmed by a positive test, whichever was earlier. Although the CIS question asks about Long Covid symptoms persisting for at least four weeks from infection, for this analysis we used a longer 12-week threshold, consistent with the UK clinical case definition of post-COVID-19 syndrome [1] and the World Health Organisation’s definition of post COVID-19 condition [22]. We therefore excluded any follow-up visits within 12 weeks of infection date.

### Exposures

The exposures of interest were first and second vaccinations of an adenovirus vector (Oxford/AstraZeneca, ChAdOx1 nCoV-19 [AZD1222]) or mRNA (Pfizer/BioNTech, BNT162b2; Moderna, mRNA-1273) COVID-19 vaccine. For each vaccine dose, we estimated the associated change in outcomes using a binary variable to indicate whether participants had received each dose at each follow-up visit; and a variable equal to the number of days since receiving each dose at each follow-up visit to estimate post-vaccination changes in the outcome trajectory (set to 0 for visits before receiving each dose). This specification implies that any change in the odds of Long Covid occurs instantly following vaccination, but in reality, this may take place over several days or weeks.

### Outcomes

The primary outcome at each visit was Long Covid of any severity. Secondary outcomes comprised Long Covid resulting in activity limitation (day-to-day activities limited “a little” or “a lot” versus “not at all” or no Long Covid); the 10 individual symptoms that were most commonly reported over the follow-up period; and whether the participant was experiencing more than three or five of the 21 symptoms included on the survey.

### Covariates

As well as time from infection and the exposure variables detailed above to modify the time trajectory of Long Covid, we adjusted for covariates hypothesised to be related to vaccine type and timing [23] and the probability of experiencing Long Covid symptoms [2]: age; sex; white or non-white ethnicity; region/country; area deprivation quintile group; health status; whether a patient-facing health or social care worker; whether hospitalised with acute COVID-19; and calendar time of infection. Specifications of these covariates can be found in Supplementary Table 3.

### Statistical analysis

We compared covariates between participants receiving adenovirus vector and mRNA vaccines using means and proportions for continuous and categorical variables, respectively. Standardized differences >10% indicated large differences [24].

Associations between exposures and outcomes were estimated using an individual-level interrupted time series approach. For each outcome, we included all exposures and covariates in a binary logistic regression model and estimated robust (clustered) standard errors to account for intra-participant correlation due to having repeated measures. We explored various specifications for modelling time since infection, finding that a linear time trend minimised the Bayesian Information Criterion for the primary outcome.

We explored heterogeneity in associations between vaccination and Long Covid by interacting all four exposure variables (change in level and slope after each dose) with each of: age group (18 to <30 years, 30 to <40 years, 40 to <50 years, 50 years to <60 years, ≥60 years); sex; white or non-white ethnicity; area deprivation quintile group; health status; hospitalisation with acute COVID-19; vaccine type (adenovirus vector or mRNA); and duration from infection to first vaccination (modelled as a restricted cubic spline). For each outcome, statistically significant interactions were identified at the 5% level after performing Holm-Bonferroni and Benjamini-Yekutieli corrections to p-values to account for multiple comparisons across exposures and modifiers. All statistical analyses were performed using R version 3.6.

### Sensitivity analyses

We restricted the sample firstly to participants with at least one observation before and after each vaccination, and secondly to those with at least three observations after each vaccination. We omitted follow-up visits within the first week after each vaccination, which may have been influenced by post-vaccine side effects. We added CIS participants who remained unvaccinated by their last follow-up visit during the study period (who were excluded from the main analysis). We excluded participants infected before the start of the second wave on 11 September 2020 [25], as mass testing for SARS-CoV-2 was largely unavailable in the first wave and so these infections were likely to have been more severe than the majority included in the analysis. Finally, we reset the infection date for 2.5% of participants where this was determined by when the participant first thought they had COVID-19 (later confirmed by a positive test) that was >14 days before a positive swab (the estimated maximum incubation period [26]). These participants may have been reinfected, but only their second infection was validated by means of a positive test, so the infection date was moved forward to the date of this test.

### Patient and public involvement

NAA has lived experience of Long Covid. Although we did not directly involve patients and the public more broadly, the study design was informed by views expressed by patient representatives in monthly meetings attended by DA (the Department of Health and Social Care’s Long Covid ministerial roundtable, NHS England’s Long Covid national taskforce).

## Results

### Description of the study sample

Of 323,685 CIS participants aged 18 to 69 years with at least one visit between 3 February and 5 September 2021, 28,356 had test-confirmed SARS-CoV-2 at least 12 weeks before their final visit and had been vaccinated post-infection, and were therefore included in analysis (Supplementary Figure 1).

Median time to the final follow-up visit was 169 (interquartile range [IQR]: 141 to 185) days from first visit, and 267 (219 to 431) days from first infection. By design, all study participants received their first vaccination by 5 September 2021, 12,971 (45.7%) after 3 February; 23,753 (83.8%) participants were double vaccinated by 5 September 2021, 20,335 (71.7%) receiving their second dose after 3 February. Supplementary Table 4 shows vaccination status during follow-up by age and health status (two of the main vaccination prioritisation determinants). Participants had a median of 4 (IQR: 2 to 5) visits after their first dose and, among those double-vaccinated, 2 (1 to 3) visits after their second dose.

At last visit, the mean age of participants was 46 years (standard deviation [SD] 14 years), 55.6% were female, and 88.7% were white (Table 1). Compared with participants receiving an adenovirus vector vaccine, those mRNA vaccinated were on average younger (mean 40 versus 51 years), and more likely to be of non-white ethnicity (13.7% versus 9.4%), resident in London (27.0% versus 22.4%) or Northern Ireland (3.3% versus 1.5%), and a patient-facing health or social care worker (17.1% versus 6.4%).

**Table 1.** Characteristics of study participants at their final follow-up visit, stratified by vaccine type. Notes: mRNA: messenger ribonucleic acid; SD: standard deviation. The study sample size did not permit disaggregation of ethnicity beyond white and non-white groups. Area deprivation was based on the English Indices of Deprivation 2019, the Welsh Index of Multiple Deprivation 2019, the Scottish Index of Multiple Deprivation 2020, and the Northern Ireland Multiple Deprivation Measure 2017. Health conditions were self-reported rather than clinically diagnosed based on the survey question: “Do you have any physical or mental health conditions or illnesses lasting or expected to last 12 months or more (excluding any long-lasting COVID-19 symptoms)?” Hospitalisation with acute COVID-19 was self-reported rather than derived from medical records.

| Characteristic | Category | Full sample<br>(*n* = 28,356) | mRNA vaccinated<br>(*n* = 12,859) | Vector vaccinated<br>(*n* = 15,497) | Standardized<br>difference (%) |
| --- | --- | --- | --- | --- | --- |
| Time since infection (days), mean (SD) |  | 308.9 (129.0) | 314.9 (132.9) | 304.0 (125.4) | 8.4 |
| Time since first vaccination (days), mean (SD) |  | 130.7 (55.9) | 118.0 (67.8) | 141.2 (40.7) | -41.5 |
| Age (years), mean (SD) |  | 45.9 (13.6) | 40.1 (14.1) | 50.7 (11.1) | -83.4 |
| Sex, *n* (%) | Male | 12,596 (44.4) | 5,466 (42.5) | 7,130 (46.0) | -7.1 |
|  | Female | 15,760 (55.6) | 7,393 (57.5) | 8,367 (54.0) | 7.1 |
| Ethnic group, *n* (%) | White | 25,141 (88.7) | 11,097 (86.3) | 14,044 (90.6) | -13.6 |
|  | Non-white | 3,215 (11.3) | 1,762 (13.7) | 1,453 (9.4) | 13.6 |
| Region or country, *n* (%) | North East England | 1,133 (4.0) | 524 (4.1) | 609 (3.9) | 0.7 |
|  | North West England | 3,990 (14.1) | 1,774 (13.8) | 2,216 (14.3) | -1.4 |
|  | Yorkshire and the Humber | 2,430 (8.6) | 1,028 (8.0) | 1,402 (9.0) | -3.8 |
|  | East Midlands | 1,755 (6.2) | 710 (5.5) | 1,045 (6.7) | -5.1 |
|  | West Midlands | 2,204 (7.8) | 917 (7.1) | 1,287 (8.3) | -4.4 |
|  | East of England | 2,447 (8.6) | 1,044 (8.1) | 1,403 (9.1) | -3.3 |
|  | London | 6,942 (24.5) | 3,470 (27.0) | 3,472 (22.4) | 10.6 |
|  | South East England | 2,919 (10.3) | 1,208 (9.4) | 1,711 (11.0) | -5.4 |
|  | South West England | 1,276 (4.5) | 580 (4.5) | 696 (4.5) | 0.1 |
|  | Northern Ireland | 657 (2.3) | 421 (3.3) | 236 (1.5) | 11.5 |
|  | Scotland | 1,376 (4.9) | 620 (4.8) | 756 (4.9) | -0.3 |
|  | Wales | 1,227 (4.3) | 563 (4.4) | 664 (4.3) | 0.5 |
| Area deprivation quintile group, *n* (%) | 1 (most deprived) | 3,825 (13.5) | 1,779 (13.8) | 2,046 (13.2) | 1.8 |
|  | 2 | 5,392 (19.0) | 2,671 (20.8) | 2,721 (17.6) | 8.2 |
|  | 3 | 5,857 (20.7) | 2,633 (20.5) | 3,224 (20.8) | -0.8 |
|  | 4 | 6,474 (22.8) | 2,914 (22.7) | 3,560 (23.0) | -0.7 |
|  | 5 (least deprived) | 6,808 (24.0) | 2,862 (22.3) | 3,946 (25.5) | -7.5 |
| Patient-facing health or social care worker, *n* (%) | Yes | 3,190 (11.2) | 2,198 (17.1) | 992 (6.4) | 33.7 |
| Health conditions, *n* (%) | Yes | 3,851 (13.6) | 1,531 (11.9) | 2,320 (15.0) | -9.0 |
| Hospitalisation with acute COVID-19, *n* (%) | Yes | 900 (3.2) | 359 (2.8) | 541 (3.5) | -4.0 |

### Long Covid trajectories before and after vaccination

Long Covid symptoms of any severity were reported by 6,729 participants (23.7%) at least once during follow-up. Before vaccination, the odds of experiencing Long Covid decreased by 0.3% (−0.9% to +0.2%) per week from infection (Table 2). First vaccination was associated with an initial 12.8% decrease (95% CI: −18.6% to −6.6%) in the odds, followed by an increase of 0.3% (−0.6% to +1.2%) per week until receiving the second dose. Second vaccination was associated with an initial 8.8% decrease (−14.1% to −3.1%) in the odds, followed by a decrease of 0.8% (−1.2% to −0.4%) per week.

**Table 2.** Estimated time trajectories of Long Covid from infection, and changes in trajectories following COVID-19 vaccination. Notes: CI: confidence interval; SE: standard error. Estimates and standard errors are on the logit scale. Odds ratios for ‘time since first/second vaccination’ represent modification of the time trajectory. Estimates and odds ratios are adjusted for age, sex, white or non-white ethnicity, region/country, area deprivation quintile group, health status, whether a patient-facing health or social care worker, whether hospitalised with acute COVID-19, and calendar time of infection.

| Outcome | Variable | Estimate | SE | P-value | Odds ratio (95% CI) |
| --- | --- | --- | --- | --- | --- |
| Long Covid of any severity | Time trajectory (per week) | -0.003 | 0.003 | 0.25 | 0.997 (0.991 to 1.002) |
|  | First vaccination (change in level) | -0.137 | 0.035 | <0.001 | 0.872 (0.814 to 0.934) |
|  | Second vaccination (change in level) | -0.092 | 0.031 | 0.003 | 0.912 (0.859 to 0.969) |
|  | Time since first vaccination (per week) | 0.006 | 0.005 | 0.21 | 1.006 (0.996 to 1.016) |
|  | Time since second vaccination (per week) | -0.011 | 0.005 | 0.03 | 0.989 (0.979 to 0.999) |
| Activity-limiting Long Covid | Time trajectory (per week) | 0.003 | 0.004 | 0.44 | 1.003 (0.996 to 1.010) |
|  | First vaccination (change in level) | -0.131 | 0.044 | 0.003 | 0.877 (0.805 to 0.955) |
|  | Second vaccination (change in level) | -0.096 | 0.038 | 0.01 | 0.909 (0.844 to 0.979) |
|  | Time since first vaccination (per week) | 0.006 | 0.006 | 0.35 | 1.006 (0.994 to 1.018) |
|  | Time since second vaccination (per week) | -0.013 | 0.006 | 0.03 | 0.987 (0.976 to 0.998) |

Long Covid resulting in activity limitation was reported by 4,747 participants (16.7%) at least once during follow-up. First vaccination was associated with an initial 12.3% decrease (−19.5% to −4.5%) in the odds of activity-limiting Long Covid, followed by an increase of 0.9% (−0.2% to +1.9%) per week until receiving the second dose. Second vaccination was associated with an initial 9.1% decrease (−15.6% to −2.1%) in the odds, followed by a decrease of 0.5% (−1.0% to +0.05%) per week.

To illustrate the impact of each vaccination, Figure 1 shows the estimated probability of reporting Long Covid for a hypothetical participant receiving their first vaccination 24 weeks after infection and their second dose 12 weeks later. Sensitivity analyses (Supplementary Figures 2a-h) were generally consistent with the main results. However, there was stronger evidence of a change to an increasing trend in Long Covid between first and second vaccinations when restricting the sample to participants who received their first dose during the follow-up period 3 February to 5 September 2021 (p<0.001 for Long Covid of any severity, p=0.007 for activity-limiting Long Covid).

**Figure 1.**
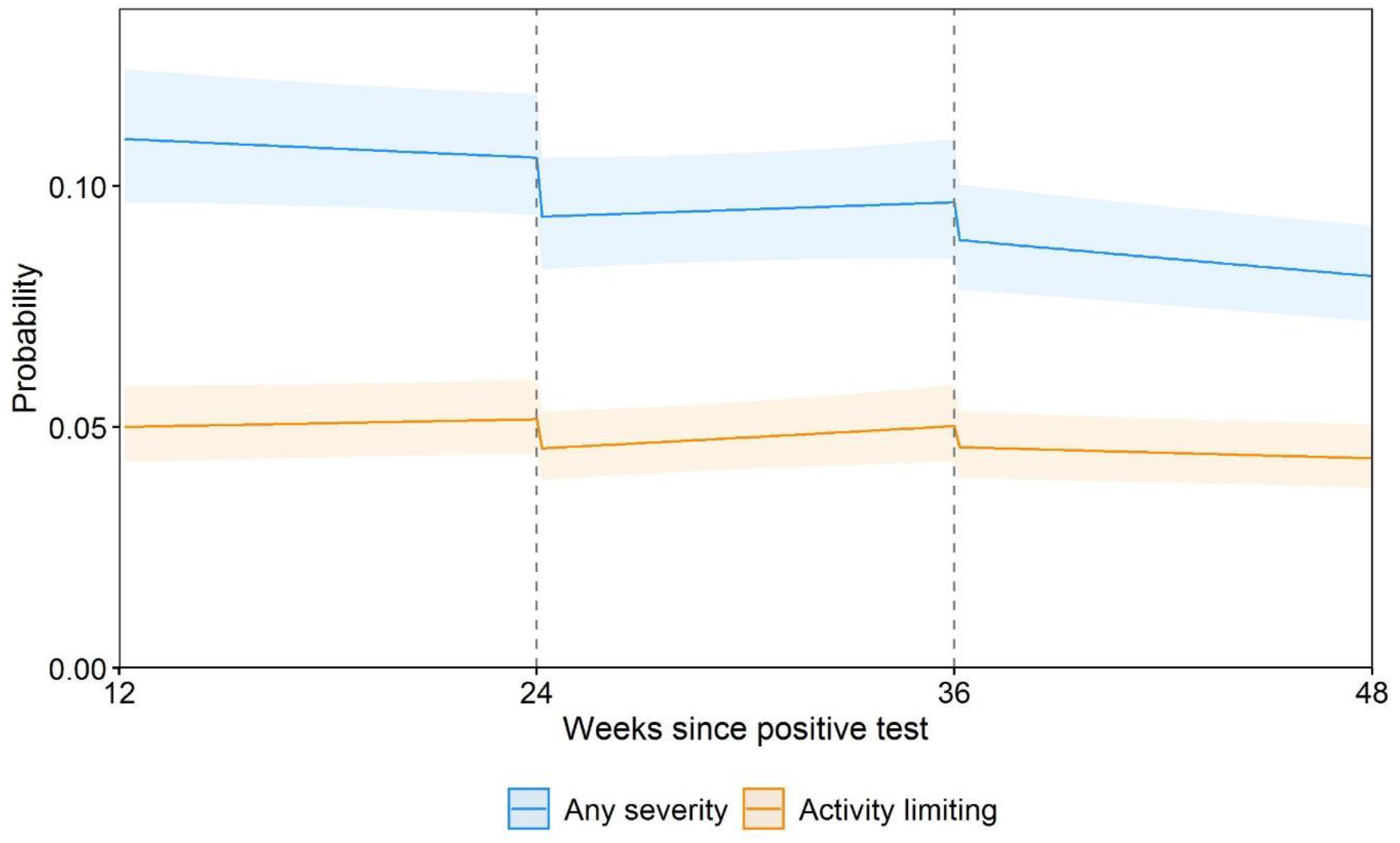
Modelled probabilities of Long Covid for a hypothetical study participant who received their first vaccination 24 weeks after infection and their second vaccination 12 weeks later. Notes: Probabilities are shown for a participant of approximately mean age (50 years) and in the modal group for other covariates (female, white, living in London, in an area in the least deprived quintile group, not a patient-facing health or social care worker, no pre-existing health conditions, not hospitalised at the acute phase of infection, and infected on 7 September 2020). While the estimated probabilities are specific to this profile, the proportional changes in probabilities after vaccination do not vary across characteristics and can therefore be generalised to other profiles. Dashed lines indicate the timing of vaccination. Shaded areas are 95% confidence intervals.

### Heterogeneity by vaccine type, duration since infection, and participant characteristics

There was no statistical evidence of differences in post-vaccination Long Covid trajectories between participants receiving adenovirus vector and mRNA vaccines (Table 3, Figure 2). Numerically, vaccination was associated with an initial 14.9% decrease (−21.8% to −7.5%) in the odds of Long Covid following first adenovirus vector vaccination, and an initial 8.9% decrease (−18.2% to +1.4%) following first mRNA vaccination. Decreases in the odds after second vaccination were numerically similar between vaccine types, at 8.7% (−15.4% to - 1.4%) for adenovirus vector and 8.9% (−17.6% to +0.7%) for mRNA.

**Figure 2.**
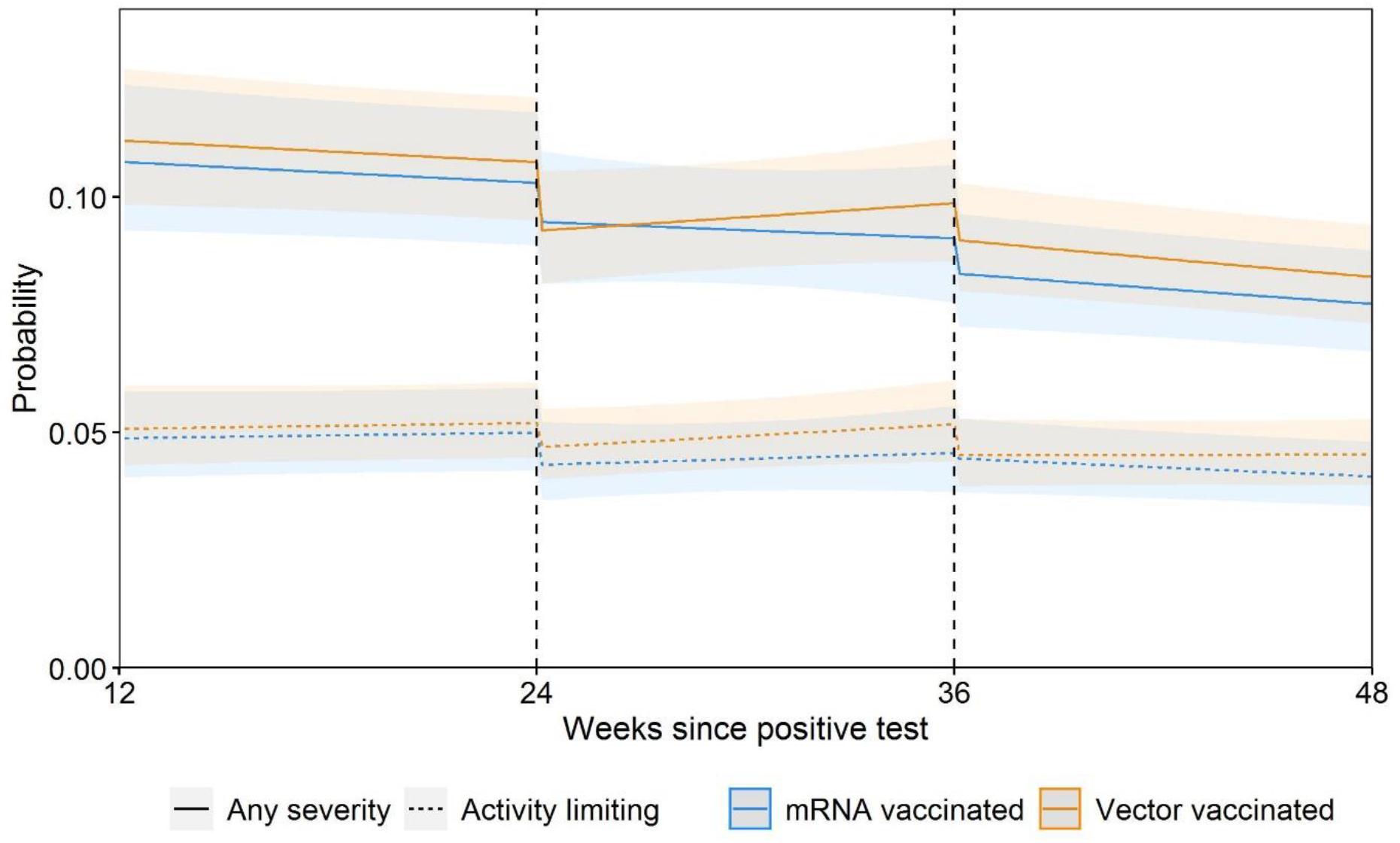
Modelled probabilities of Long Covid for hypothetical study participants who received their first adenovirus vector or mRNA vaccination 24 weeks after infection and their second vaccination 12 weeks later. Notes: mRNA: messenger ribonucleic acid. Probabilities are shown for a participant of approximately mean age (50 years) and in the modal group for other covariates (female, white, living in London, in an area in the least deprived quintile group, not a patient-facing health or social care worker, no pre-existing health conditions, not hospitalised at the acute phase of infection, and infected on 7 September 2020). While the estimated probabilities are specific to this profile, the proportional changes in probabilities after vaccination do not vary across characteristics and can therefore be generalised to other profiles. Dashed lines indicate the timing of vaccination. Shaded areas are 95% confidence intervals.

**Table 3.** Estimated time trajectories of Long Covid from infection, and changes in trajectories following COVID-19 vaccination, moderated by vaccine type. Notes: CI: confidence interval; mRNA: messenger ribonucleic acid; SE: standard error. Estimates and standard errors are on the logit scale. Odds ratios for ‘time since first/second vaccination’ represent modification of the time trajectory. Odds ratios for ‘first/second vaccination interacted with type’ represent modification of the change in level after first/second vaccination by vaccine type. Odds ratios for ‘time since first/second vaccination interacted with type’ represent modification of the time trajectory, modified by vaccine type. Estimates and odds ratios are adjusted for age, sex, white or non-white ethnicity, region/country, area deprivation quintile group, health status, whether a patient-facing health or social care worker, whether hospitalised with acute COVID-19, and calendar time of infection.

| Outcome | Variable | Estimate | SE | P-value | Odds ratio (95% CI) |
| --- | --- | --- | --- | --- | --- |
| Long Covid of any severity | Time trajectory (per week) | -0.004 | 0.003 | 0.19 | 0.996 (0.990 to 1.002) |
|  | First vaccination (change in level) | -0.093 | 0.055 | 0.09 | 0.911 (0.818 to 1.014) |
|  | Second vaccination (change in level) | -0.093 | 0.051 | 0.07 | 0.911 (0.824 to 1.007) |
|  | Time since first vaccination (per week) | 0.000 | 0.008 | 0.95 | 1.000 (0.985 to 1.016) |
|  | Time since second vaccination (per week) | -0.004 | 0.008 | 0.65 | 0.996 (0.980 to 1.013) |
|  | Vaccine type: adenovirus vector (versus mRNA) | 0.046 | 0.055 | 0.40 | 1.048 (0.941 to 1.166) |
|  | First vaccination interacted with type | -0.069 | 0.067 | 0.31 | 0.934 (0.818 to 1.066) |
|  | Second vaccination interacted with type | 0.002 | 0.064 | 0.97 | 1.002 (0.883 to 1.137) |
|  | Time since first vaccination interacted with type | 0.009 | 0.009 | 0.33 | 1.009 (0.991 to 1.028) |
|  | Time since second vaccination interacted with type | -0.010 | 0.010 | 0.33 | 0.990 (0.970 to 1.010) |
| Activity-limiting Long Covid | Time trajectory (per week) | 0.002 | 0.004 | 0.56 | 1.002 (0.995 to 1.009) |
|  | First vaccination (change in level) | -0.154 | 0.070 | 0.03 | 0.857 (0.747 to 0.984) |
|  | Second vaccination (change in level) | -0.026 | 0.064 | 0.68 | 0.974 (0.860 to 1.103) |
|  | Time since first vaccination (per week) | 0.003 | 0.010 | 0.77 | 1.003 (0.984 to 1.022) |
|  | Time since second vaccination (per week) | -0.013 | 0.010 | 0.21 | 0.987 (0.968 to 1.007) |
|  | Vaccine type: adenovirus vector (versus mRNA) | 0.042 | 0.069 | 0.54 | 1.043 (0.911 to 1.195) |
|  | First vaccination interacted with type | 0.045 | 0.087 | 0.60 | 1.046 (0.883 to 1.240) |
|  | Second vaccination interacted with type | -0.116 | 0.080 | 0.15 | 0.890 (0.761 to 1.041) |
|  | Time since first vaccination interacted with type | 0.004 | 0.012 | 0.75 | 1.004 (0.981 to 1.027) |
|  | Time since second vaccination interacted with type | 0.004 | 0.013 | 0.73 | 1.004 (0.980 to 1.029) |

The odds of Long Covid after first vaccination numerically decreased with duration from infection, with estimated decreases of 24.8%, 16.5%, and 4.8% for participants first vaccinated 9, 12, and 15 months after infection (Supplementary Figures 3a-b). However, duration from infection to first vaccination was not a statistically significant moderator of the vaccination-Long Covid relationship (Supplementary Tables 5a to 5d).

There was no statistical evidence of differences in post-vaccination Long Covid trends according to socio-demographic characteristics (age, sex, ethnic group, area deprivation) or health-related factors (self-reported health status not related to COVID-19, whether hospitalised with acute COVID-19) (Supplementary Tables 5a to 5d).

### Trajectories of individual symptoms

The odds of experiencing most symptoms, as well as more than three or five symptoms together, initially numerically decreased after each vaccination (Figure 3). After first vaccination, the largest numerical decreases were observed for loss of smell (−12.5%, - 21.5% to −2.5%), loss of taste (−9.2%, −19.8% to +2.7%), and trouble sleeping (−8.8%, - 19.4% to +3.3%). After second vaccination, the largest numerical decreases were observed for fatigue (−9.7%, −16.5% to −2.4%), headache (−9.0%, −18.1% to +1.0%), and trouble sleeping (−9.0%, −18.2% to +1.2%).

**Figure 3.**
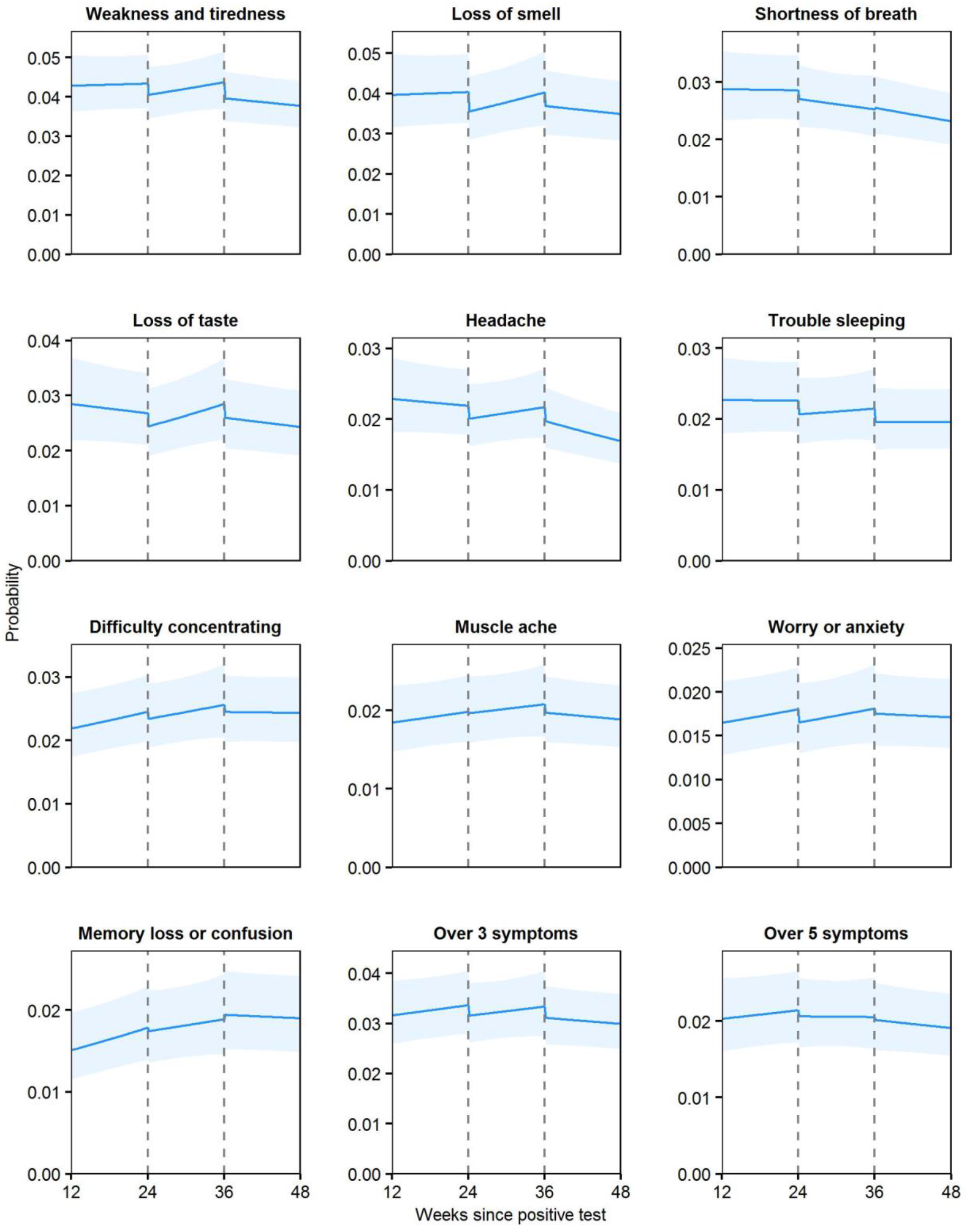
Modelled probabilities of individual Long Covid symptoms for a hypothetical study participant who received their first vaccination 24 weeks after infection and their second vaccination 12 weeks later. Notes: Top 10 most frequently reported symptoms ordered by modelled probability at 12 weeks post-infection. Probabilities are shown for a participant of approximately mean age (50 years) and in the modal group for other covariates (female, white, living in London, in an area in the least deprived quintile group, not a patient-facing health or social care worker, no pre-existing health conditions, not hospitalised at the acute phase of infection, and infected on 7 September 2020). While the estimated probabilities are specific to this profile, the proportional changes in probabilities after vaccination do not vary across characteristics and can therefore be generalised to other profiles. Dashed lines indicate the timing of vaccination. Shaded areas are 95% confidence intervals.

Similar to Long Covid overall, the odds of experiencing most individual symptoms, and more than three or five symptoms together, numerically decreased after the first vaccination. Trends were generally upwards between the first and second vaccinations, with most returning to a declining or flat trend after the second dose. However, lack of statistical power meant that for most symptoms, the data were compatible with both initial increases and decreases, and with both upward and downward trends, in the likelihood of experiencing symptoms after each vaccination (Supplementary Table 6).

## Discussion

In this community-based study of adults aged 18 to 69 years infected with SARS-CoV-2 prior to vaccination, we found that the odds of experiencing Long Covid symptoms that had persisted for at least 12 weeks fell by an average of 13% after receiving a first COVID-19 vaccination. However, it is unclear from the data whether the improvement was sustained until receiving the second vaccination. Receiving a second vaccination was associated with a further 9% decrease in the odds of Long Covid, and there was statistical evidence of a sustained improvement after this, at least over the median follow-up time of 67 days. Similar findings were obtained when focussing on Long Covid severe enough to result in functional impairment.

We found no statistical evidence of heterogeneity in the associations between vaccination and Long Covid symptoms according to vaccine type, duration from infection to first vaccination, socio-demographic characteristics including age, sex, ethnicity, and area deprivation, self-reported health status, and whether hospitalised with acute COVID-19. However, this observational study was unlikely to have been sufficiently powered to detect these associations, particularly given the multiplicity of testing, and absence of evidence does not necessarily imply evidence of absence.

### Findings in context

Our results substantially add to existing evidence on the epidemiology of Long Covid after vaccination. While previous studies are generally coherent in terms of their findings, ours exploits a considerably larger sample drawn at random from the community population, so the results are likely to be more precise and generalizable.

A study of 44 vaccinated patients and 22 unvaccinated controls previously hospitalised with COVID-19 in the UK, which inevitably had limited power to detect clinically relevant effects, found no evidence of vaccination being associated with worsening of Long Covid symptoms or quality of life [17]. A study of 455 self-selected participants in France found reduced symptom burden and double the rate of remission at 120 days post-vaccination compared with unvaccinated controls [18]. A non-controlled study of 900 social media users found that over half had experienced an improvement in symptoms after vaccination while just 7% reported a deterioration [19].

COVID-19 vaccination effectively reduces rates of infection [11–12] and transmission [13]. Evidence also suggests that Long Covid incidence is reduced in those infected after vaccination; in a study of 906 mobile phone app users, the odds of having symptoms ≥28 days post-infection was approximately halved in fully vaccinated participants versus unvaccinated controls [16]. Together with our results, these findings suggest that COVID-19 vaccination may reduce the population prevalence of Long Covid by reducing the risk of continuing to experience persistent symptoms in those who already have them when vaccinated; developing persistent symptoms following breakthrough infections; being infected in the first place; and transmitting the virus following infection.

Our principal finding, of a decrease in the likelihood of experiencing Long Covid symptoms after receiving a second vaccination, supports hypothesised biological mechanisms. People with Long Covid experiencing dysregulation of the immune system may benefit from vaccine-induced diversion of autoimmune processes, while any residual viral reservoir may be destroyed by the antibody response [27]. However, whether this is a long-lasting ‘reset’ of the immune system remains to be established.

The symptom trajectory following the initial fall after first vaccination was unclear, being compatible with both increasing and decreasing odds of Long Covid over time. However, there was evidence of an increasing trend when the sample was restricted to participants vaccinated during the follow-up period. Relapsing symptoms are common in Long Covid [3–5] and persistent symptoms are associated with weak antibody response [28], so it is possible that receiving a first dose alone is insufficient for sustained improvement in some people.

### Strengths and limitations

With 28,356 adults in our sample, this is the largest study to date internationally on Long Covid and COVID-19 vaccination, and the first to investigate post-vaccine symptom trajectories. The main strength of the study is its use of the CIS, a large survey of approximately half a million people from the community population of the UK with longitudinal follow-up. Random sampling from address lists mitigates against selection bias, while the prospective design means that survey responses are not subject to outcome recall bias (such as participants overestimating the duration of previously experienced symptoms). All CIS participants are swabbed for SARS-CoV-2 at every follow-up visit, irrespective of symptoms, so our study includes asymptomatic as well symptomatic infections.

The study also has limitations. Its observational nature means that causality cannot be inferred, and placebo and side effects of vaccination may have contributed to our findings; however, estimates were robust to excluding follow-up visits within the first week of each vaccination, suggesting that the impact of these effects is likely to be small. Although we adjusted for a wide range of potential confounders, unmeasured factors, such as those related to take-up of a second vaccination, may remain.

The observed changes after vaccination could be related to the relapsing-remitting nature of symptoms experienced by many people living with Long Covid [3–5] rather than a causal effect of the vaccine. Future analysis should consider differing patterns of illness, including quantification of the frequency and duration of symptom-free periods after vaccination. Although all infections were test-confirmed, Long Covid status was self-reported and we did not have data on formal clinical diagnoses, so we cannot exclude some participants’ symptoms being caused by a medical condition other than SARS-CoV-2 infection.

It is possible that the average improvement in Long Covid symptoms and functional impact may wane with time, and longer-term follow-up is required to establish whether the estimated changes after second vaccination are sustained. Follow-up after receipt of a booster dose, now widely available in the UK adult population, is also required. The study sample was restricted to participants aged 18 to 69 years, so our findings may not generalize to children or older adults, nor may they apply to people who had not received a vaccine by 5 September 2021, in particular those who are vaccine-hesitant because of their Long Covid symptoms. However, our results were insensitive to whether unvaccinated participants were included in the study sample.

### Conclusions

In summary, we found that COVID-19 vaccination is associated with a decrease in the likelihood of continuing to experience Long Covid symptoms in adults aged 18 to 69 years, and this appeared to be sustained after the second dose. Our results suggest that vaccination of people previously infected may be associated with a reduction in the burden of Long Covid on population health, at least in the first few months following vaccination. Further research is required to evaluate the long-term relationship between vaccination and Long Covid, and understand the biological mechanisms underpinning any improvements in symptoms following vaccination. Such research may contribute to the development of therapeutics for Long Covid.

## Supporting information

STROBE checklist

## Data Availability

De-identified study data are available to accredited researchers in the ONS Secure Research Service (SRS) under part 5, chapter 5 of the Digital Economy Act 2017. For further information about accreditation, contact or visit: ons.gov.uk/aboutus/whatwedo/statistics/requestingstatistics/approvedresearcherscheme

## Acknowledgements

KK and FZ are supported by the National Institute for Health Research (NIHR) Applied Research Collaboration East Midlands (ARC EM) and the NIHR Leicester Biomedical Research Centre (BRC). KBP and ASW are supported by the NIHR Health Protection Research Unit in Healthcare Associated Infections and Antimicrobial Resistance (NIHR200915), a partnership between the UK Health Security Agency (UKHSA) and the University of Oxford. KBP is also supported by the Huo Family Foundation. ASW is also supported by the NIHR Oxford Biomedical Research Centre and is an NIHR Senior Investigator. NAA has lived experience of Long Covid and is a co-investigator on the NIHR-funded STIMULATE-ICP study. The views expressed are those of the authors and are not necessarily those of the National Health Service, the NIHR, the Department of Health and Social Care, or the UK Health Security Agency.

## Footnotes

### Contributors

DA, KBP, MG, VN, NAA and ASW conceptualised and designed the study. DA and CB prepared the study data and performed the statistical analysis. All authors contributed to interpretation of the results. DA and CB were responsible for the first draft of the manuscript. All authors contributed to critical revision of the manuscript. All authors approved the final manuscript.

The lead author (the manuscript’s guarantor) affirms that the manuscript is an honest, accurate, and transparent account of the study being reported; that no important aspects of the study have been omitted; and that any discrepancies from the study as originally planned (and, if relevant, registered) have been explained.

### Ethical approval

Ethical approval for this study was obtained from the National Statistician’s Data Ethics Advisory Committee (NSDEC(20)12). The CIS received ethical approval from the South Central Berkshire B Research Ethics Committee (20/SC/0195).

### Data sharing

Dissemination to participants and related patient and public communities: The use of de-identified data precludes direct dissemination to participants. For the purpose of open access, the authors have applied a Creative Commons Attribution (CC BY) licence to any Author Accepted Manuscript version arising

---

**Supplementary Table 1.** Number and percentage of households invited to participate in the Coronavirus (COVID-19) Infection Survey who subsequently enrolled, by country and phase. Notes: The initial invitation phase started on 26 April 2020 in England, 29 June 2020 in Wales, and 26 July 2020 in Northern Ireland. The extension period started on 31 May 2020 in England. Sampling from AddressBase started on 13 July 2020 in England, 5 October 2020 in Wales, and 14 September 2020 in Scotland. Enrolment rates as of 22 October 2021, taken from the technical dataset accompanying the Coronavirus (COVID-19) Infection Survey: https://www.ons.gov.uk/peoplepopulationandcommunity/healthandsocialcare/conditionsanddiseases/d atasets/covid19infectionsurveytechnicaldata

| Phase | England | Wales | Northern Ireland | Scotland |
| --- | --- | --- | --- | --- |
| Initial invitation | 10,266 (51%) | 7,017 (40%) | 7,084 (43%) | N/A |
| Extension period | 39,345 (43%) | N/A | N/A | N/A |
| AddressBase | 167,890 (12%) | 7,013 (14%) | N/A | 22,131 (13%) |

**Supplementary Table 2.** Long Covid symptoms included on the Coronavirus (COVID-19) Infection Survey. Notes: All participants who responded positively to the survey question “Would you describe yourself as having’long COVID’, that is, you are still experiencing symptoms more than 4 weeks after you first had COVID-19, that are not explained by something else?” were then asked “Do you have any of the following symptoms as part of your experience of long COVID? Please include any pre-existing symptoms which long COVID has made worse (answer Yes or No for each one).”

| Symptom |
| --- |
| 1. Fever |
| 2. Headache |
| 3. Muscle ache |
| 4. Weakness/tiredness |
| 5. Nausea/vomiting |
| 6. Abdominal pain |
| 7. Diarrhoea |
| 8. Loss of appetite |
| 9. Loss of taste |
| 10. Loss of smell |
| 11. Sore throat |
| 12. Cough |
| 13. Shortness of breath |
| 14. Chest pain |
| 15. Palpitations |
| 16. Vertigo/dizziness |
| 17. Worry/anxiety |
| 18. Low mood/not enjoying anything |
| 19. Trouble sleeping |
| 20. Memory loss or confusion |
| 21. Difficulty concentrating |

**Supplementary Table 3.**
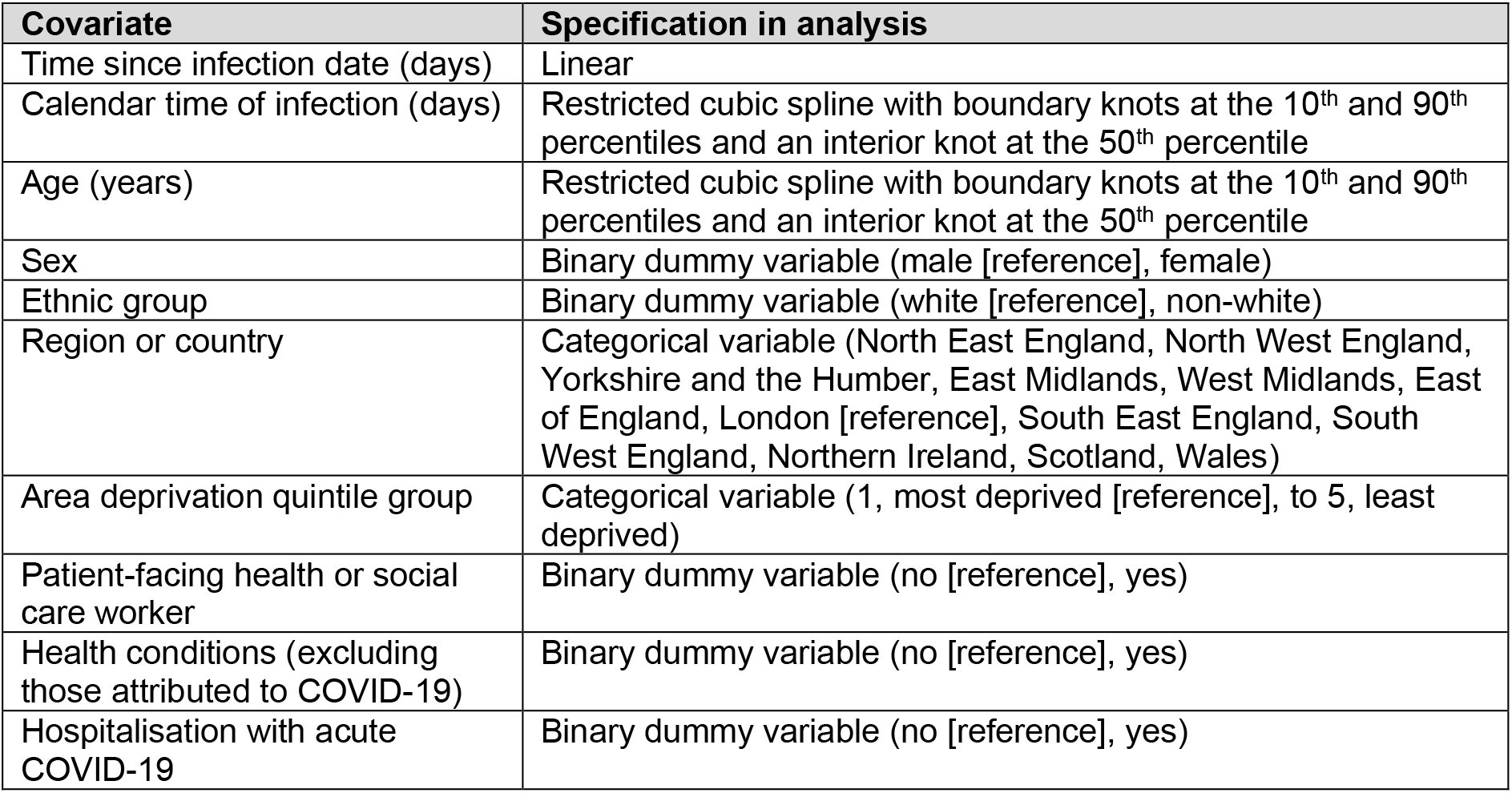
Description of covariates included in the analysis. Notes: Calendar time of infection was calculated as the number of days from 24 January 2020 (when the first SARS-CoV-2 cases were reported in the UK) and the infection date. The study sample size did not permit disaggregation of ethnicity beyond white and non-white groups. Area deprivation was based on the English Indices of Deprivation 2019, the Welsh Index of Multiple Deprivation 2019, the Scottish Index of Multiple Deprivation 2020, and the Northern Ireland Multiple Deprivation Measure 2017. Health conditions were self-reported rather than clinically diagnosed based on the survey question: “Do you have any physical or mental health conditions or illnesses lasting or expected to last 12 months or more (excluding any long-lasting COVID-19 symptoms)?” Hospitalisation with acute COVID-19 was self-reported rather than derived from medical records.

**Supplementary Table 4.** Number and percentage of included participants who received their first and second vaccinations during (rather than before) the follow-up period, stratified by age group and health conditions at last follow-up visit. Notes: The study sample included only people aged under 70 years at their first visit during the follow-up period, and a small number of participants turned 70 before their last visit, hence the upper boundary of the final age group is 70 rather than 69 years. Health conditions were self-reported rather than clinically diagnosed based on the survey question: “Do you have any physical or mental health conditions or illnesses lasting or expected to last 12 months or more (excluding any long-lasting COVID-19 symptoms)?”

| Age group | Health conditions | Total | Received first vaccination during follow-up | Received second vaccination during follow-up |
| --- | --- | --- | --- | --- |
| 18 to 29 years | No | 3,789 | 2,367 (62.5%) | 1,666 (44.0%) |
|  | Yes | 385 | 197 (51.2%) | 213 (55.3%) |
| 30 to 39 years | No | 4,902 | 2,921 (59.6%) | 3,264 (66.6%) |
|  | Yes | 533 | 238 (44.7%) | 359 (67.4%) |
| 40 to 49 years | No | 5,675 | 2,923 (51.5%) | 4,325 (76.2%) |
|  | Yes | 699 | 291 (41.6%) | 534 (76.4%) |
| 50 to 59 years | No | 5,829 | 2,275 (39.0%) | 4,719 (81.0%) |
|  | Yes | 1,112 | 394 (35.4%) | 888 (79.9%) |
| 60 to 70 years | No | 4,310 | 1,088 (25.2%) | 3,507 (81.4%) |
|  | Yes | 1,122 | 277 (24.7%) | 860 (76.7%) |

**Supplementary Table 5a.**
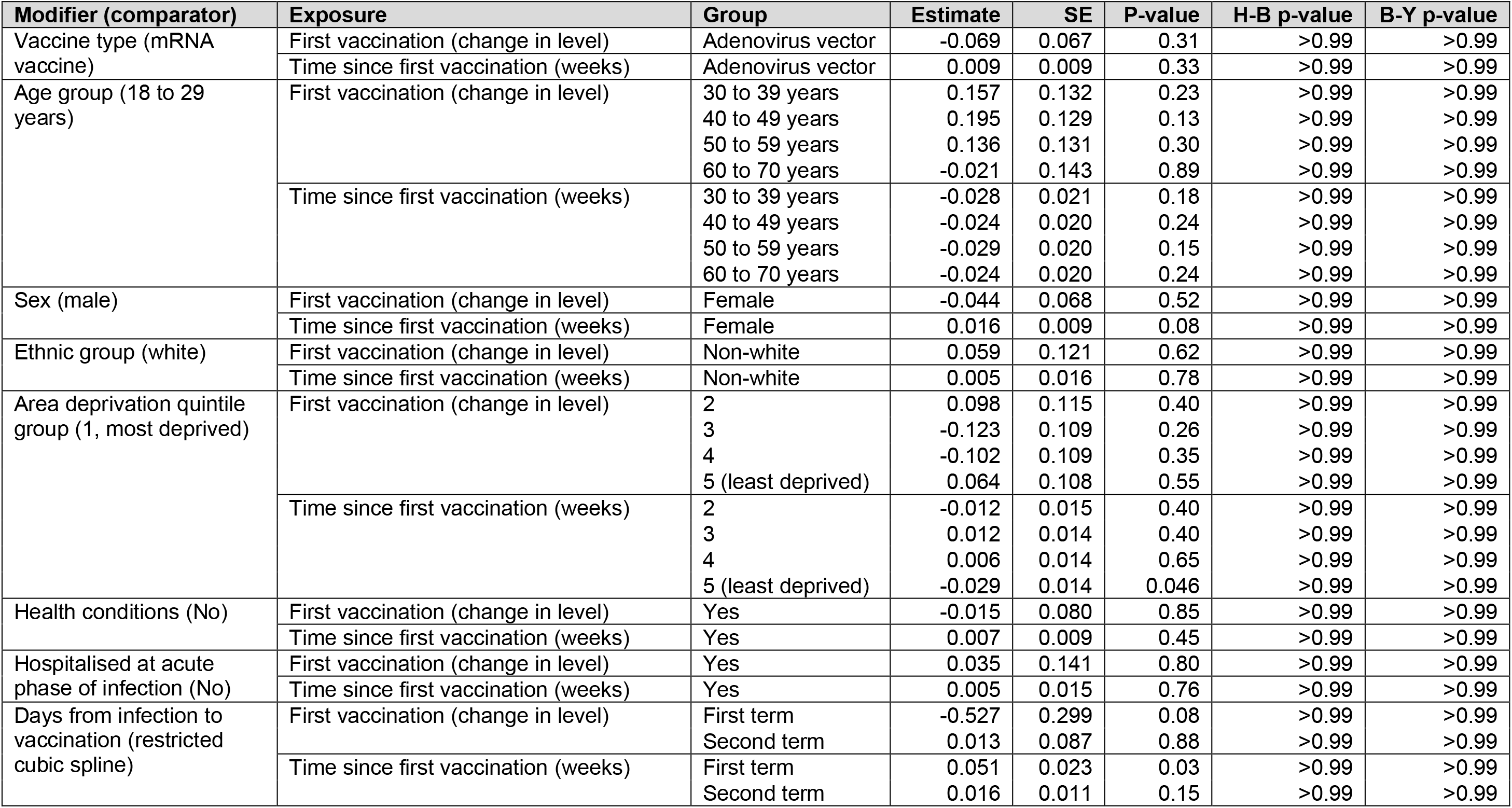
Interactions between vaccination and personal characteristics: Long Covid of any severity, first vaccination. Notes: B-Y: Benjamini-Yekutieli; H-B: Holm-Bonferroni; mRNA: messenger ribonucleic acid; SE: standard error.

| Modifier (comparator) | Exposure | Group | Estimate | SE | P-value | H-B p-value | B-Y p-value |
| --- | --- | --- | --- | --- | --- | --- | --- |
| Vaccine type (mRNA vaccine) | First vaccination (change in level) | Adenovirus vector | -0.069 | 0.067 | 0.31 | >0.99 | >0.99 |
|  | Time since first vaccination (weeks) | Adenovirus vector | 0.009 | 0.009 | 0.33 | >0.99 | >0.99 |
| Age group (18 to 29 years) | First vaccination (change in level) | 30 to 39 years | 0.157 | 0.132 | 0.23 | >0.99 | >0.99 |
|  |  | 40 to 49 years | 0.195 | 0.129 | 0.13 | >0.99 | >0.99 |
|  |  | 50 to 59 years | 0.136 | 0.131 | 0.30 | >0.99 | >0.99 |
|  |  | 60 to 70 years | -0.021 | 0.143 | 0.89 | >0.99 | >0.99 |
|  | Time since first vaccination (weeks) | 30 to 39 years | -0.028 | 0.021 | 0.18 | >0.99 | >0.99 |
|  |  | 40 to 49 years | -0.024 | 0.020 | 0.24 | >0.99 | >0.99 |
|  |  | 50 to 59 years | -0.029 | 0.020 | 0.15 | >0.99 | >0.99 |
|  |  | 60 to 70 years | -0.024 | 0.020 | 0.24 | >0.99 | >0.99 |
| Sex (male) | First vaccination (change in level) | Female | -0.044 | 0.068 | 0.52 | >0.99 | >0.99 |
|  | Time since first vaccination (weeks) | Female | 0.016 | 0.009 | 0.08 | >0.99 | >0.99 |
| Ethnic group (white) | First vaccination (change in level) | Non-white | 0.059 | 0.121 | 0.62 | >0.99 | >0.99 |
|  | Time since first vaccination (weeks) | Non-white | 0.005 | 0.016 | 0.78 | >0.99 | >0.99 |
| Area deprivation quintile group (1, most deprived) | First vaccination (change in level) | 2 | 0.098 | 0.115 | 0.40 | >0.99 | >0.99 |
|  |  | 3 | -0.123 | 0.109 | 0.26 | >0.99 | >0.99 |
|  |  | 4 | -0.102 | 0.109 | 0.35 | >0.99 | >0.99 |
|  |  | 5 (least deprived) | 0.064 | 0.108 | 0.55 | >0.99 | >0.99 |
|  | Time since first vaccination (weeks) | 2 | -0.012 | 0.015 | 0.40 | >0.99 | >0.99 |
|  |  | 3 | 0.012 | 0.014 | 0.40 | >0.99 | >0.99 |
|  |  | 4 | 0.006 | 0.014 | 0.65 | >0.99 | >0.99 |
|  |  | 5 (least deprived) | -0.029 | 0.014 | 0.046 | >0.99 | >0.99 |
| Health conditions (No) | First vaccination (change in level) | Yes | -0.015 | 0.080 | 0.85 | >0.99 | >0.99 |
|  | Time since first vaccination (weeks) | Yes | 0.007 | 0.009 | 0.45 | >0.99 | >0.99 |
| Hospitalised at acute phase of infection (No) | First vaccination (change in level) | Yes | 0.035 | 0.141 | 0.80 | >0.99 | >0.99 |
|  | Time since first vaccination (weeks) | Yes | 0.005 | 0.015 | 0.76 | >0.99 | >0.99 |
| Days from infection to vaccination (restricted cubic spline) | First vaccination (change in level) | First term | -0.527 | 0.299 | 0.08 | >0.99 | >0.99 |
|  |  | Second term | 0.013 | 0.087 | 0.88 | >0.99 | >0.99 |
|  | Time since first vaccination (weeks) | First term | 0.051 | 0.023 | 0.03 | >0.99 | >0.99 |
|  |  | Second term | 0.016 | 0.011 | 0.15 | >0.99 | >0.99 |

**Supplementary Table 5b.** Interactions between vaccination and personal characteristics: Long Covid of any severity, second vaccination. Notes: B-Y: Benjamini-Yekutieli; H-B: Holm-Bonferroni; mRNA: messenger ribonucleic acid; SE: standard error.

| Modifier (comparator) | Exposure | Group | Estimate | SE | P-value | H-B p-value | B-Y p-value |
| --- | --- | --- | --- | --- | --- | --- | --- |
| Vaccine type (mRNA vaccine) | Second vaccination (change in level) | Adenovirus vector | 0.002 | 0.064 | 0.97 | >0.99 | >0.99 |
|  | Time since second vaccination (weeks) | Adenovirus vector | -0.010 | 0.010 | 0.33 | >0.99 | >0.99 |
| Age group (18 to 29 years) | Second vaccination (change in level) | 30 to 39 years | 0.182 | 0.145 | 0.21 | >0.99 | >0.99 |
|  |  | 40 to 49 years | 0.206 | 0.136 | 0.13 | >0.99 | >0.99 |
|  |  | 50 to 59 years | 0.239 | 0.134 | 0.07 | >0.99 | >0.99 |
|  |  | 60 to 70 years | 0.177 | 0.137 | 0.19 | >0.99 | >0.99 |
|  | Time since second vaccination (weeks) | 30 to 39 years | 0.041 | 0.026 | 0.12 | >0.99 | >0.99 |
|  |  | 40 to 49 years | 0.037 | 0.025 | 0.15 | >0.99 | >0.99 |
|  |  | 50 to 59 years | 0.035 | 0.025 | 0.17 | >0.99 | >0.99 |
|  |  | 60 to 70 years | 0.033 | 0.026 | 0.20 | >0.99 | >0.99 |
| Sex (male) | Second vaccination (change in level) | Female | -0.124 | 0.064 | 0.06 | >0.99 | >0.99 |
|  | Time since second vaccination (weeks) | Female | -0.015 | 0.011 | 0.16 | >0.99 | >0.99 |
| Ethnic group (white) | Second vaccination (change in level) | Non-white | -0.092 | 0.112 | 0.41 | >0.99 | >0.99 |
|  | Time since second vaccination (weeks) | Non-white | -0.004 | 0.018 | 0.81 | >0.99 | >0.99 |
| Area deprivation quintile group (1, most deprived) | Second vaccination (change in level) | 2 | 0.123 | 0.108 | 0.26 | >0.99 | >0.99 |
|  |  | 3 | 0.026 | 0.105 | 0.81 | >0.99 | >0.99 |
|  |  | 4 | 0.041 | 0.103 | 0.69 | >0.99 | >0.99 |
|  |  | 5 (least deprived) | 0.242 | 0.104 | 0.02 | >0.99 | >0.99 |
|  | Time since second vaccination (weeks) | 2 | 0.001 | 0.017 | 0.94 | >0.99 | >0.99 |
|  |  | 3 | -0.026 | 0.016 | 0.09 | >0.99 | >0.99 |
|  |  | 4 | -0.015 | 0.015 | 0.34 | >0.99 | >0.99 |
|  |  | 5 (least deprived) | 0.017 | 0.016 | 0.30 | >0.99 | >0.99 |
| Health conditions (No) | Second vaccination (change in level) | Yes | -0.049 | 0.071 | 0.49 | >0.99 | >0.99 |
|  | Time since second vaccination (weeks) | Yes | 0.004 | 0.010 | 0.67 | >0.99 | >0.99 |
| Hospitalised at acute phase of infection (No) | Second vaccination (change in level) | Yes | 0.076 | 0.113 | 0.50 | >0.99 | >0.99 |
|  | Time since second vaccination (weeks) | Yes | -0.005 | 0.017 | 0.79 | >0.99 | >0.99 |
| Days from infection to vaccination (restricted cubic spline) | Second vaccination (change in level) | First term | -0.107 | 0.142 | 0.45 | >0.99 | >0.99 |
|  |  | Second term | -0.025 | 0.085 | 0.77 | >0.99 | >0.99 |
|  | Time since second vaccination (weeks) | First term | -0.057 | 0.025 | 0.02 | >0.99 | >0.99 |
|  |  | Second term | -0.005 | 0.014 | 0.72 | >0.99 | >0.99 |

**Supplementary Table 5c.** Interactions between vaccination and personal characteristics: activity-limiting Long Covid, first vaccination. Notes: B-Y: Benjamini-Yekutieli; H-B: Holm-Bonferroni; mRNA: messenger ribonucleic acid; SE: standard error.

| Modifier (comparator) | Exposure | Group | Estimate | SE | P-value | H-B p-value | B-Y p-value |
| --- | --- | --- | --- | --- | --- | --- | --- |
| Vaccine type (mRNA vaccine) | First vaccination (change in level) | Adenovirus vector | 0.045 | 0.087 | 0.60 | >0.99 | >0.99 |
|  | Time since first vaccination (weeks) | Adenovirus vector | 0.004 | 0.012 | 0.75 | >0.99 | >0.99 |
| Age group (18 to 29 years) | First vaccination (change in level) | 30 to 39 years | 0.267 | 0.199 | 0.18 | >0.99 | >0.99 |
|  |  | 40 to 49 years | 0.306 | 0.195 | 0.12 | >0.99 | >0.99 |
|  |  | 50 to 59 years | 0.273 | 0.195 | 0.16 | >0.99 | >0.99 |
|  |  | 60 to 70 years | 0.203 | 0.206 | 0.33 | >0.99 | >0.99 |
|  | Time since first vaccination (weeks) | 30 to 39 years | -0.029 | 0.032 | 0.36 | >0.99 | >0.99 |
|  |  | 40 to 49 years | -0.024 | 0.031 | 0.45 | >0.99 | >0.99 |
|  |  | 50 to 59 years | -0.025 | 0.031 | 0.42 | >0.99 | >0.99 |
|  |  | 60 to 70 years | -0.023 | 0.031 | 0.45 | >0.99 | >0.99 |
| Sex (male) | First vaccination (change in level) | Female | -0.123 | 0.086 | 0.15 | >0.99 | >0.99 |
|  | Time since first vaccination (weeks) | Female | 0.023 | 0.011 | 0.04 | >0.99 | >0.99 |
| Ethnic group (white) | First vaccination (change in level) | Non-white | -0.030 | 0.154 | 0.85 | >0.99 | >0.99 |
|  | Time since first vaccination (weeks) | Non-white | 0.019 | 0.019 | 0.34 | >0.99 | >0.99 |
| Area deprivation quintile group (1, most deprived) | First vaccination (change in level) | 2 | 0.385 | 0.141 | 0.006 | 0.35 | 0.56 |
|  |  | 3 | 0.045 | 0.133 | 0.74 | >0.99 | >0.99 |
|  |  | 4 | 0.138 | 0.132 | 0.30 | >0.99 | >0.99 |
|  |  | 5 (least deprived) | 0.135 | 0.135 | 0.32 | >0.99 | >0.99 |
|  | Time since first vaccination (weeks) | 2 | -0.028 | 0.018 | 0.11 | >0.99 | >0.99 |
|  |  | 3 | 0.008 | 0.016 | 0.62 | >0.99 | >0.99 |
|  |  | 4 | -0.015 | 0.016 | 0.36 | >0.99 | >0.99 |
|  |  | 5 (least deprived) | -0.022 | 0.017 | 0.20 | >0.99 | >0.99 |
| Health conditions (No) | First vaccination (change in level) | Yes | -0.030 | 0.092 | 0.75 | >0.99 | >0.99 |
|  | Time since first vaccination (weeks) | Yes | -0.001 | 0.010 | 0.91 | >0.99 | >0.99 |
| Hospitalised at acute phase of infection (No) | First vaccination (change in level) | Yes | 0.135 | 0.155 | 0.38 | >0.99 | >0.99 |
|  | Time since first vaccination (weeks) | Yes | -0.015 | 0.017 | 0.38 | >0.99 | >0.99 |
| Days from infection to vaccination (restricted cubic spline) | First vaccination (change in level) | First term | -0.606 | 0.374 | 0.11 | >0.99 | >0.99 |
|  |  | Second term | -0.031 | 0.111 | 0.78 | >0.99 | >0.99 |
|  | Time since first vaccination (weeks) | First term | 0.078 | 0.028 | 0.005 | 0.30 | 0.56 |
|  |  | Second term | 0.027 | 0.013 | 0.04 | >0.99 | >0.99 |

**Supplementary Table 5d.** Interactions between vaccination and personal characteristics: activity-limiting Long Covid, second vaccination. Notes: B-Y: Benjamini-Yekutieli; H-B: Holm-Bonferroni; mRNA: messenger ribonucleic acid; SE: standard error.

| Modifier (comparator) | Exposure | Group | Estimate | SE | P-value | H-B p-value | B-Y p-value |
| --- | --- | --- | --- | --- | --- | --- | --- |
| Vaccine type (mRNA vaccine) | Second vaccination (change in level) | Adenovirus vector | -0.116 | 0.080 | 0.15 | >0.99 | >0.99 |
|  | Time since second vaccination (weeks) | Adenovirus vector | 0.004 | 0.013 | 0.73 | >0.99 | >0.99 |
| Age group (18 to 29 years) | Second vaccination (change in level) | 30 to 39 years | 0.112 | 0.198 | 0.57 | >0.99 | >0.99 |
|  |  | 40 to 49 years | 0.051 | 0.187 | 0.79 | >0.99 | >0.99 |
|  |  | 50 to 59 years | 0.104 | 0.183 | 0.57 | >0.99 | >0.99 |
|  |  | 60 to 70 years | 0.065 | 0.185 | 0.73 | >0.99 | >0.99 |
|  | Time since second vaccination (weeks) | 30 to 39 years | 0.045 | 0.041 | 0.28 | >0.99 | >0.99 |
|  |  | 40 to 49 years | 0.035 | 0.040 | 0.38 | >0.99 | >0.99 |
|  |  | 50 to 59 years | 0.029 | 0.040 | 0.47 | >0.99 | >0.99 |
|  |  | 60 to 70 years | 0.027 | 0.040 | 0.50 | >0.99 | >0.99 |
| Sex (male) | Second vaccination (change in level) | Female | -0.141 | 0.080 | 0.08 | >0.99 | >0.99 |
|  | Time since second vaccination (weeks) | Female | -0.025 | 0.013 | 0.047 | >0.99 | >0.99 |
| Ethnic group (white) | Second vaccination (change in level) | Non-white | -0.169 | 0.138 | 0.22 | >0.99 | >0.99 |
|  | Time since second vaccination (weeks) | Non-white | -0.011 | 0.021 | 0.61 | >0.99 | >0.99 |
| Area deprivation quintile group (1, most deprived) | Second vaccination (change in level) | 2 | 0.190 | 0.129 | 0.14 | >0.99 | >0.99 |
|  |  | 3 | -0.046 | 0.125 | 0.71 | >0.99 | >0.99 |
|  |  | 4 | 0.107 | 0.124 | 0.39 | >0.99 | >0.99 |
|  |  | 5 (least deprived) | 0.247 | 0.125 | 0.048 | >0.99 | >0.99 |
|  | Time since second vaccination (weeks) | 2 | 0.017 | 0.020 | 0.40 | >0.99 | >0.99 |
|  |  | 3 | -0.022 | 0.018 | 0.22 | >0.99 | >0.99 |
|  |  | 4 | 0.004 | 0.018 | 0.82 | >0.99 | >0.99 |
|  |  | 5 (least deprived) | 0.010 | 0.019 | 0.60 | >0.99 | >0.99 |
| Health conditions (No) | Second vaccination (change in level) | Yes | 0.031 | 0.080 | 0.70 | >0.99 | >0.99 |
|  | Time since second vaccination (weeks) | Yes | 0.008 | 0.011 | 0.48 | >0.99 | >0.99 |
| Hospitalised at acute phase of infection (No) | Second vaccination (change in level) | Yes | 0.087 | 0.124 | 0.48 | >0.99 | >0.99 |
|  | Time since second vaccination (weeks) | Yes | 0.014 | 0.019 | 0.45 | >0.99 | >0.99 |
| Days from infection to vaccination (restricted cubic spline) | Second vaccination (change in level) | First term | -0.074 | 0.174 | 0.67 | >0.99 | >0.99 |
|  |  | Second term | -0.080 | 0.105 | 0.44 | >0.99 | >0.99 |
|  | Time since second vaccination (weeks) | First term | -0.089 | 0.029 | 0.002 | 0.12 | 0.56 |
|  |  | Second term | -0.015 | 0.017 | 0.35 | >0.99 | >0.99 |

**Supplementary Table 6.**
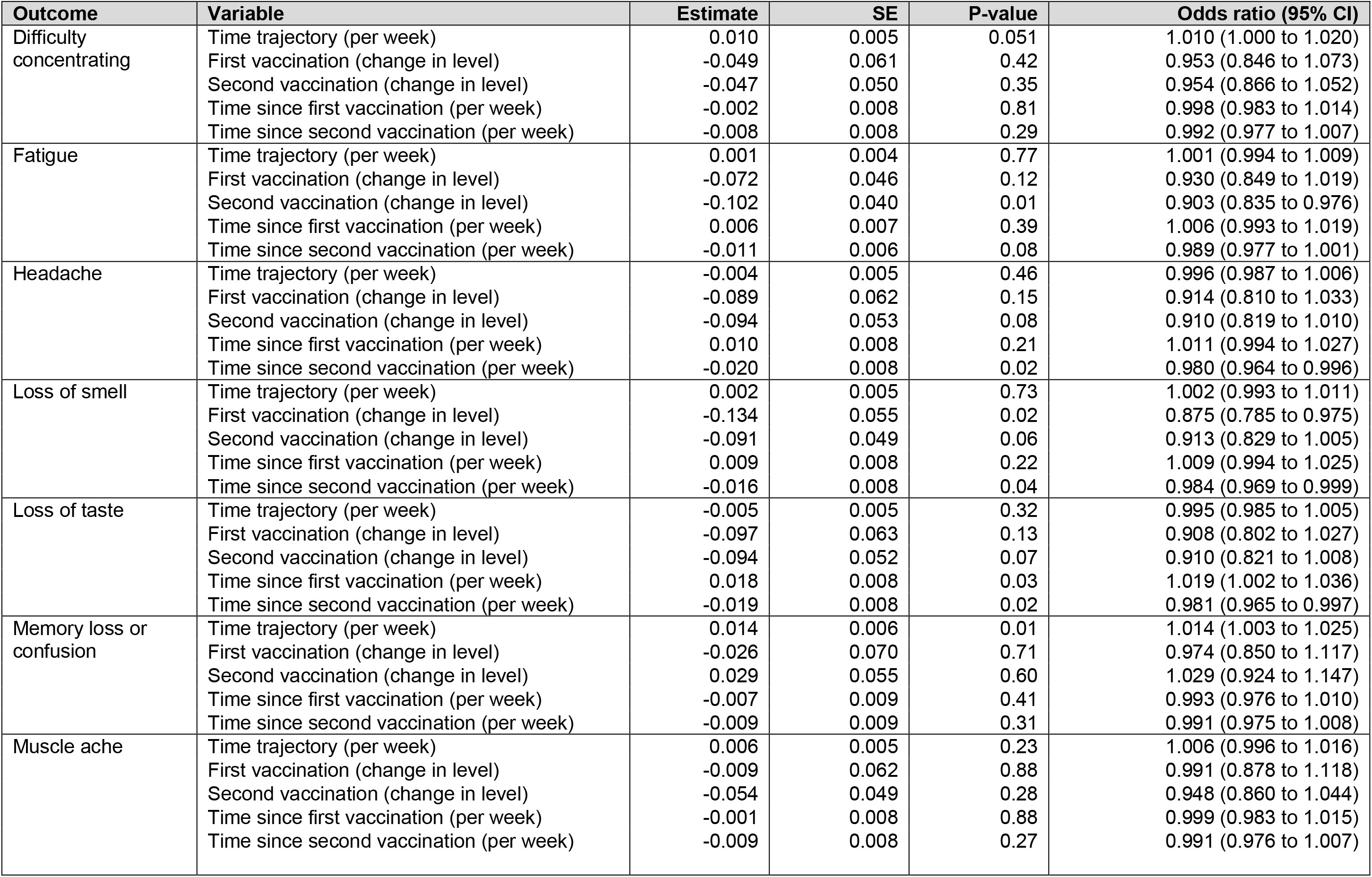
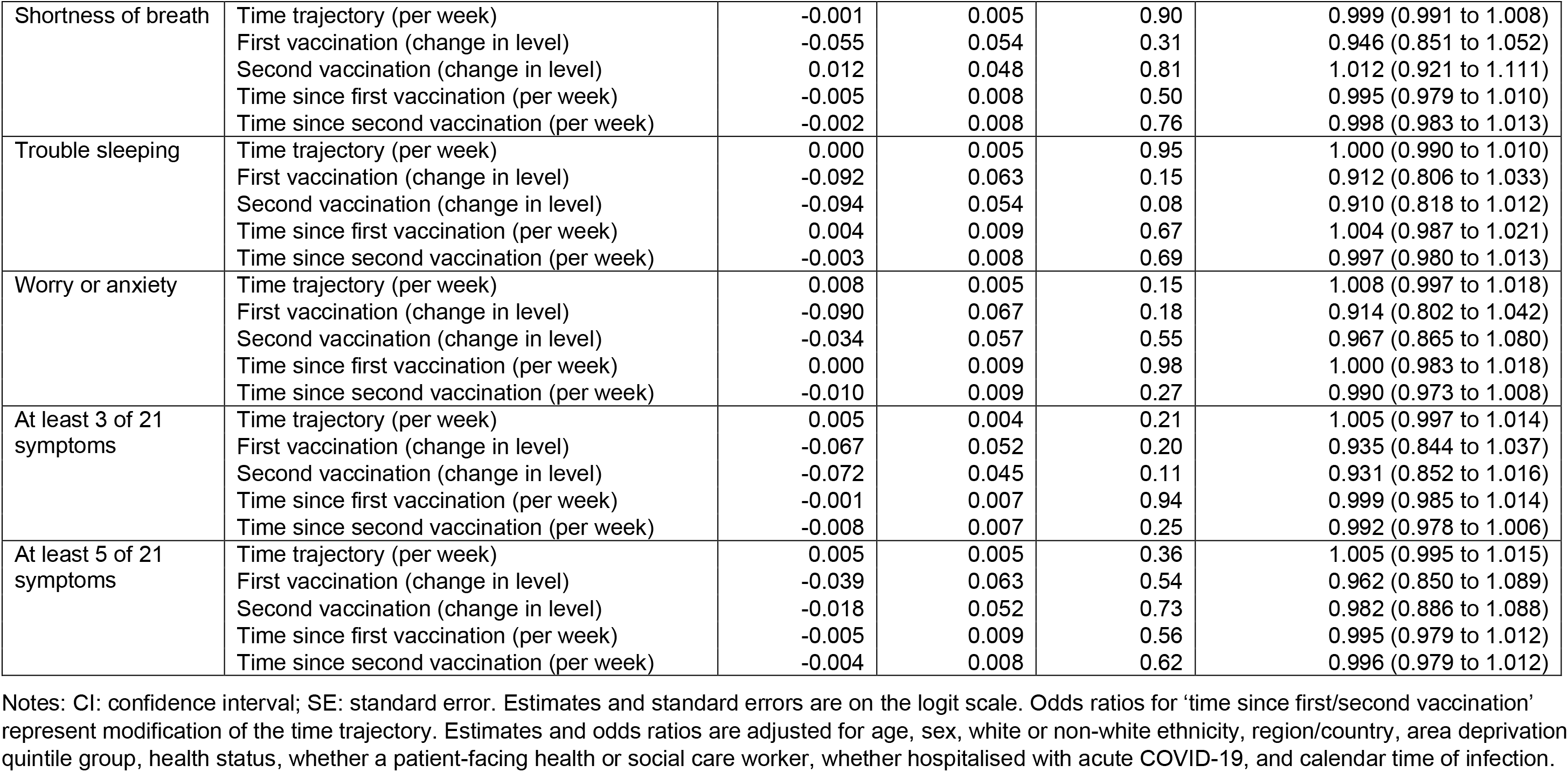
Logistic regression model output for time trends in individual Long Covid symptoms and vaccination exposure variables. Notes: CI: confidence interval; SE: standard error. Estimates and standard errors are on the logit scale. Odds ratios for ‘time since first/second vaccination’ represent modification of the time trajectory. Estimates and odds ratios are adjusted for age, sex, white or non-white ethnicity, region/country, area deprivation quintile group, health status, whether a patient-facing health or social care worker, whether hospitalised with acute COVID-19, and calendar time of infection.

**Supplementary Figure 1.**
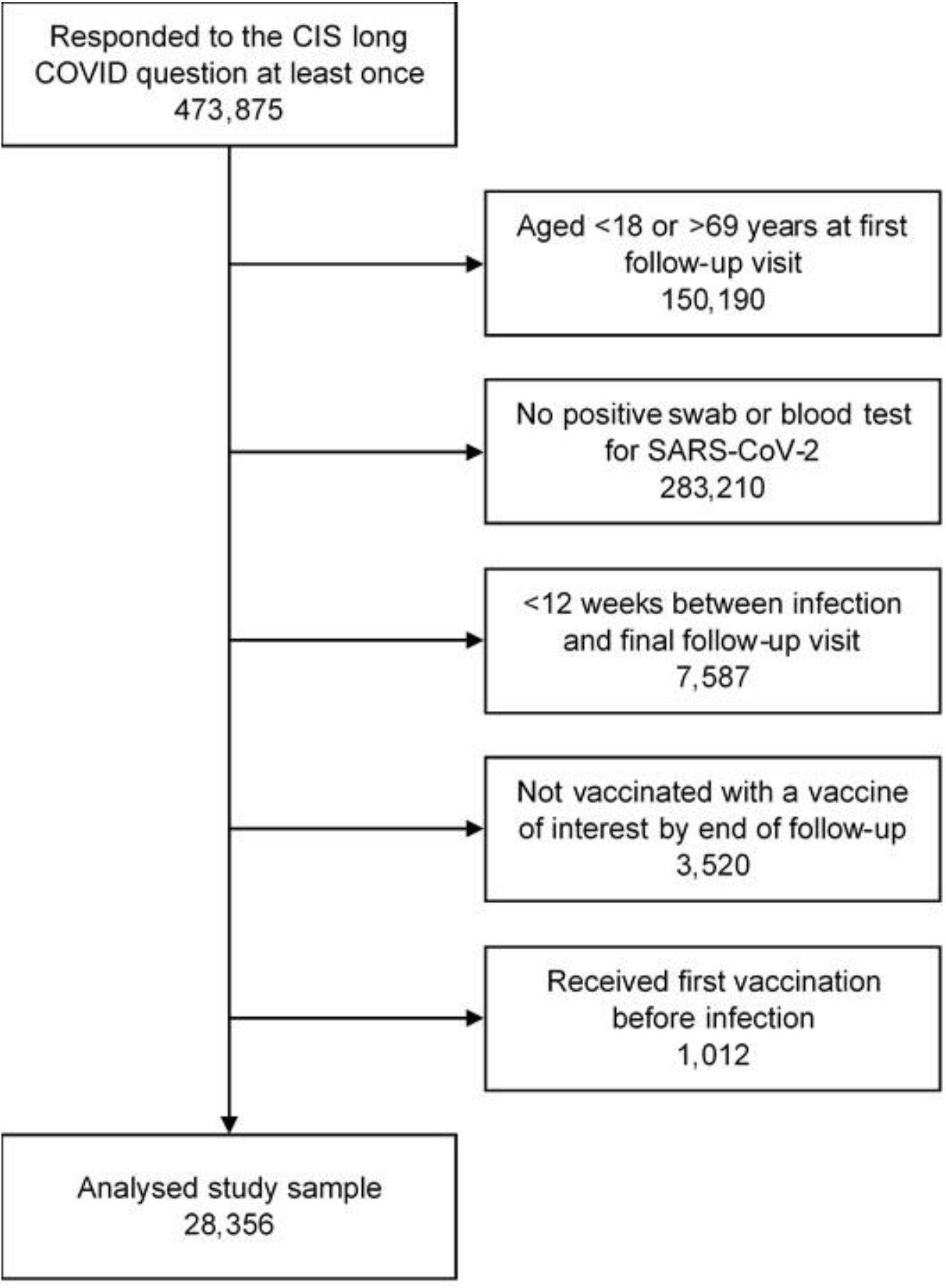
Study participant flow diagram.

**Supplementary Figure 2a.**
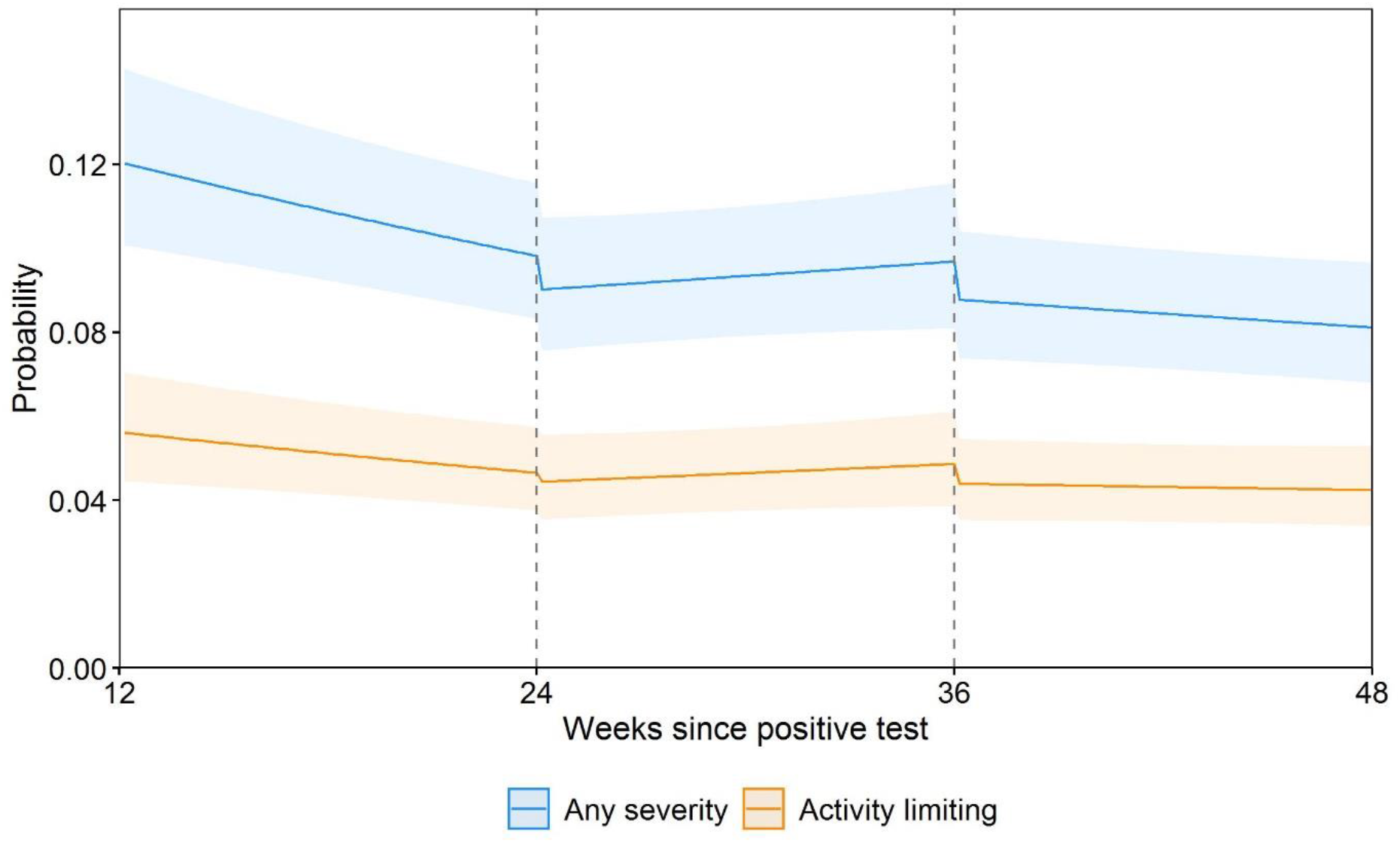
Modelled probabilities of Long Covid for a hypothetical study participant, sensitivity analysis 1: participants with at least one observation before and after their first vaccination. Notes: The sample comprised 12,971 participants with a mean age of 42 years and 10.8% with underlying health conditions (two of the main determinants of vaccination timing). Probabilities are shown for a participant aged 50 years and in the modal group for other covariates (female, white, living in London, in an area in the least deprived quintile group, not a patient-facing health or social care worker, no pre-existing health conditions, not hospitalised at the acute phase of infection, and infected on 7 September 2020). While the estimated probabilities are specific to this profile, the proportional changes in probabilities after vaccination do not vary across characteristics and can therefore be generalised to other profiles. Dashed lines indicate the timing of vaccination. Shaded areas are 95% confidence intervals.

**Supplementary Figure 2b.**
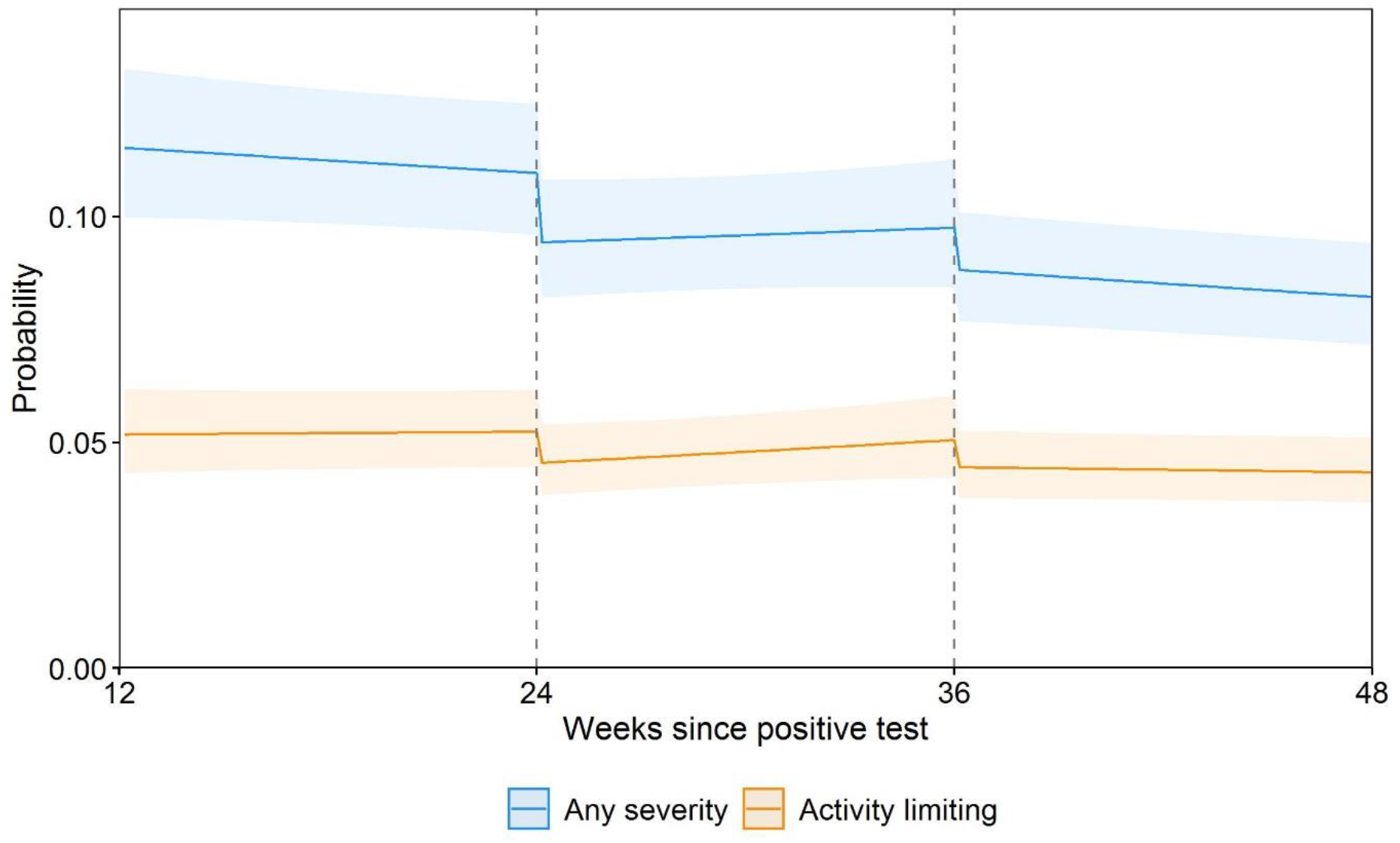
Modelled probabilities of Long Covid for a hypothetical study participant, sensitivity analysis 2: participants with at least one observation before and after their second vaccination. Notes: The sample comprises 20,335 participants with a mean age of 48 years and 14.0% with underlying health conditions (two of the main determinants of vaccination timing). Probabilities are shown for a participant aged 50 years and in the modal group for other covariates (female, white, living in London, in an area in the least deprived quintile group, not a patient-facing health or social care worker, no pre-existing health conditions, not hospitalised at the acute phase of infection, and infected on 7 September 2020). While the estimated probabilities are specific to this profile, the proportional changes in probabilities after vaccination do not vary across characteristics and can therefore be generalised to other profiles. Dashed lines indicate the timing of vaccination. Shaded areas are 95% confidence intervals.

**Supplementary Figure 2c.**
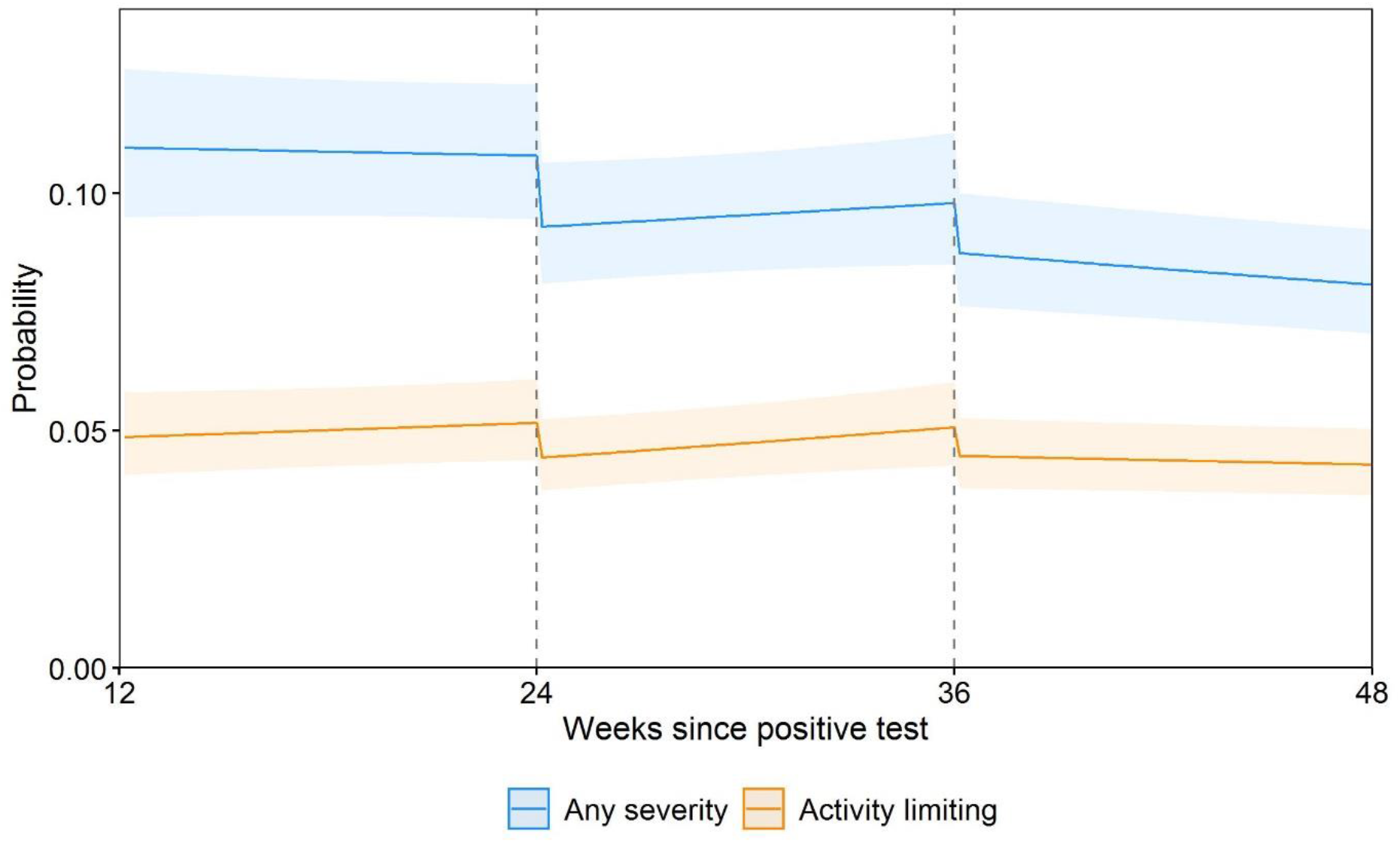
Modelled probabilities of Long Covid for a hypothetical study participant, sensitivity analysis 3: participants with at least three observations after their first vaccination. Notes: The sample comprises 20,635 participants with a mean age of 49 years and 15.1% with underlying health conditions (two of the main determinants of vaccination timing). Probabilities are shown for a participant aged 50 years and in the modal group for other covariates (female, white, living in London, in an area in the least deprived quintile group, not a patient-facing health or social care worker, no pre-existing health conditions, not hospitalised at the acute phase of infection, and infected on 7 September 2020). While the estimated probabilities are specific to this profile, the proportional changes in probabilities after vaccination do not vary across characteristics and can therefore be generalised to other profiles. Dashed lines indicate the timing of vaccination. Shaded areas are 95% confidence intervals.

**Supplementary Figure 2d.**
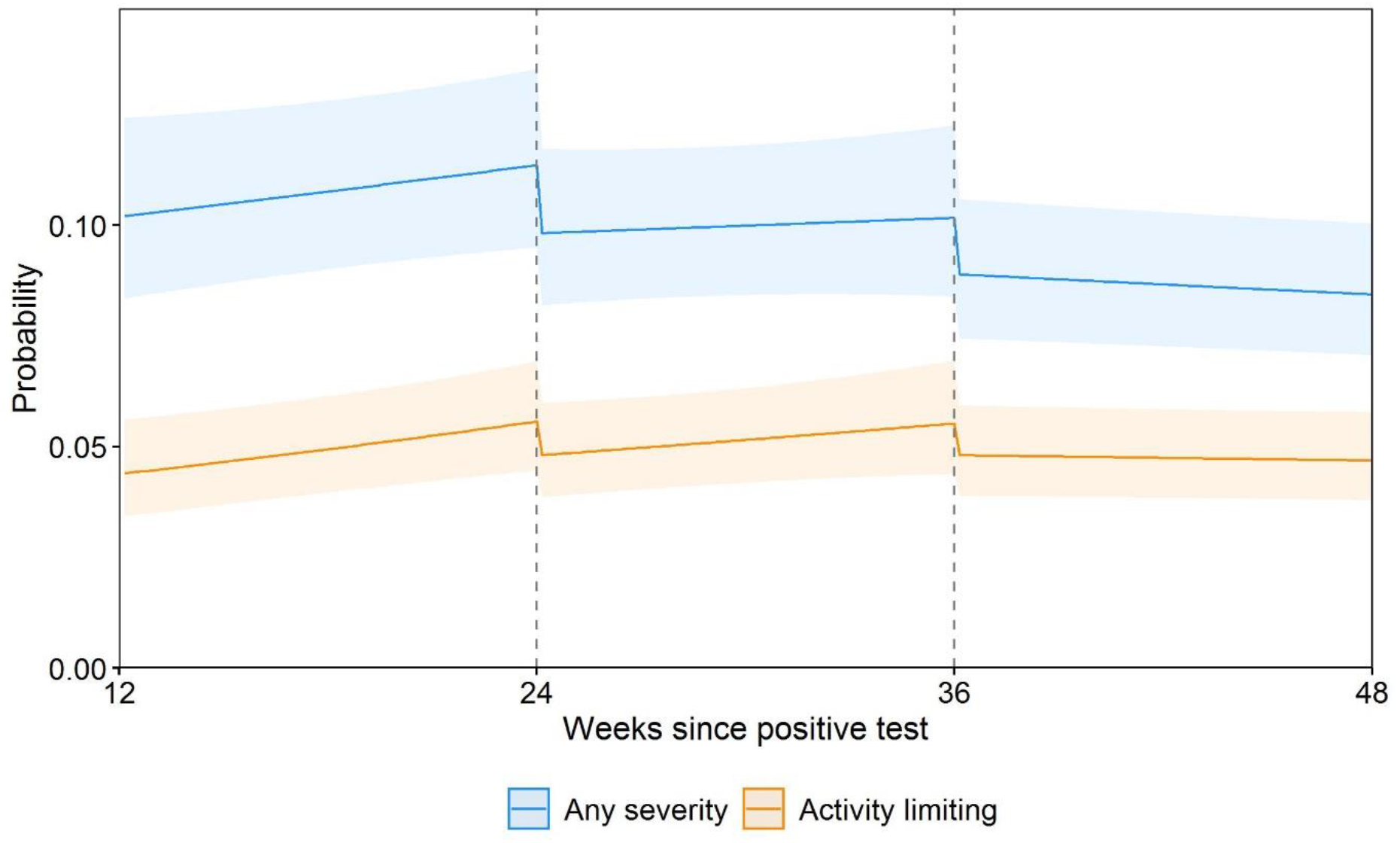
Modelled probabilities of Long Covid for a hypothetical study participant, sensitivity analysis 4: participants with at least three observations after their second vaccination. Notes: The sample comprises 12,288 participants with a mean age of 52 years and 18.0% with underlying health conditions (two of the main determinants of vaccination timing). Probabilities are shown for a participant aged 50 years and in the modal group for other covariates (female, white, living in London, in an area in the least deprived quintile group, not a patient-facing health or social care worker, no pre-existing health conditions, not hospitalised at the acute phase of infection, and infected on 7 September 2020). While the estimated probabilities are specific to this profile, the proportional changes in probabilities after vaccination do not vary across characteristics and can therefore be generalised to other profiles. Dashed lines indicate the timing of vaccination. Shaded areas are 95% confidence intervals.

**Supplementary Figure 2e.**
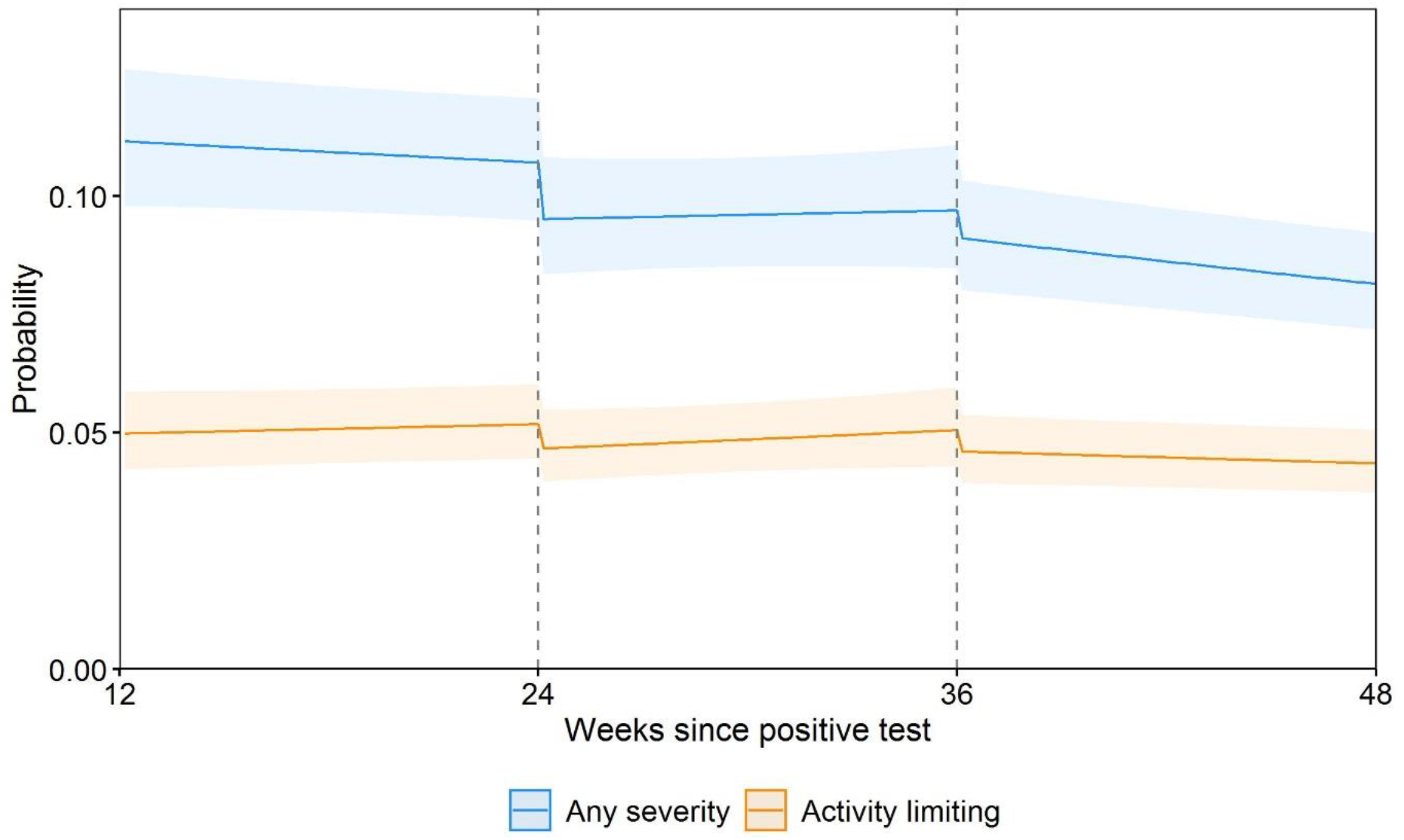
Modelled probabilities of Long Covid for a hypothetical study participant, sensitivity analysis 5: omitting follow-up visits within the first week after each vaccination. Notes: This analysis was based on 120,077 (89.6%) of the 133,965 follow-up visits used in the main analysis. Probabilities are shown for a participant aged 50 years and in the modal group for other covariates (female, white, living in London, in an area in the least deprived quintile group, not a patient-facing health or social care worker, no pre-existing health conditions, not hospitalised at the acute phase of infection, and infected on 7 September 2020). While the estimated probabilities are specific to this profile, the proportional changes in probabilities after vaccination do not vary across characteristics and can therefore be generalised to other profiles. Dashed lines indicate the timing of vaccination. Shaded areas are 95% confidence intervals.

**Supplementary Figure 2f.**
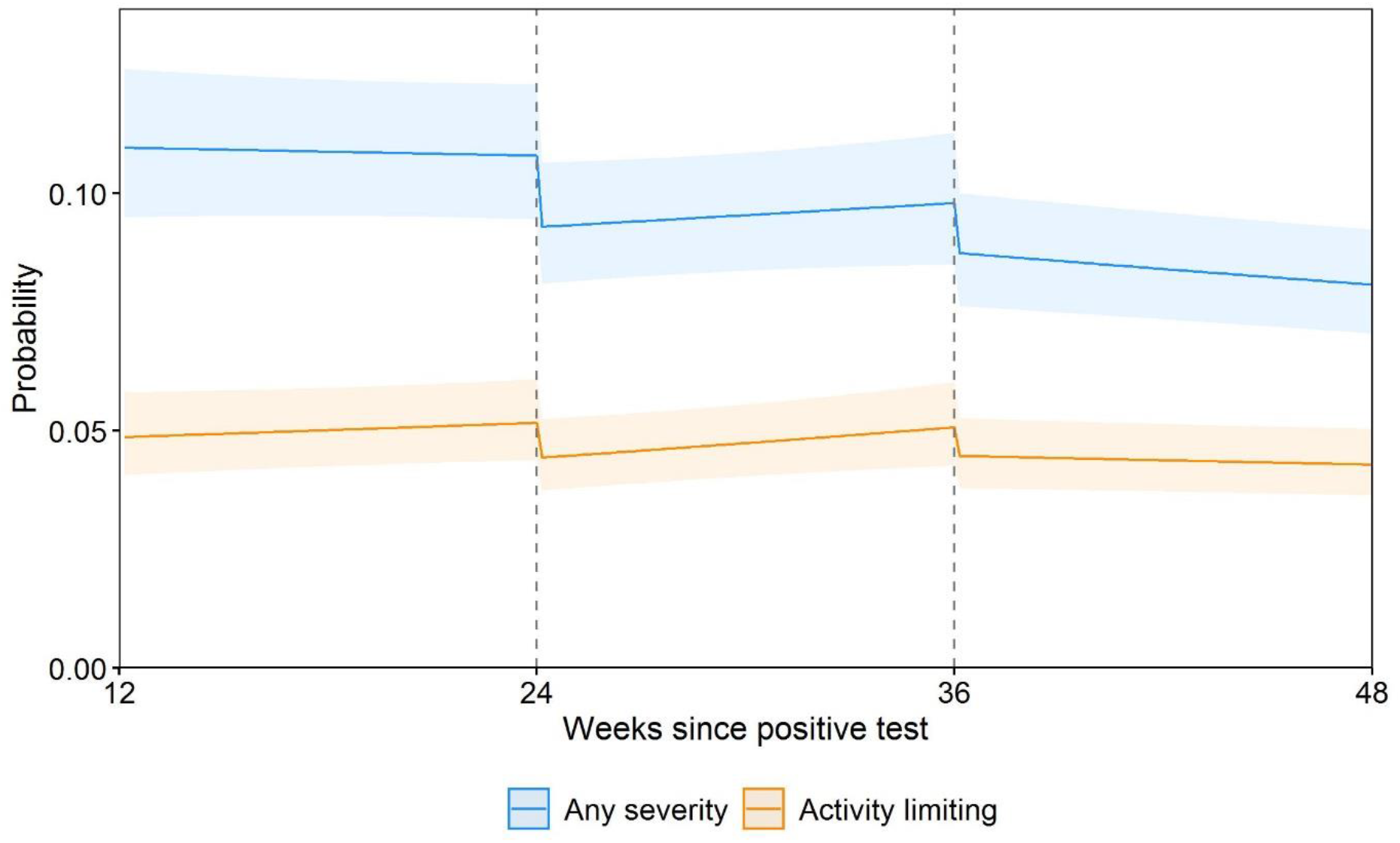
Modelled probabilities of Long Covid for a hypothetical study participant, sensitivity analysis 6: including participants who remained unvaccinated by their last follow-up visit during the study period. Notes: This analysis was based on 31,663 participants (compared with 28,356 in the main analysis). Probabilities are shown for a participant aged 50 years and in the modal group for other covariates (female, white, living in London, in an area in the least deprived quintile group, not a patient-facing health or social care worker, no pre-existing health conditions, not hospitalised at the acute phase of infection, and infected on 7 September 2020). While the estimated probabilities are specific to this profile, the proportional changes in probabilities after vaccination do not vary across characteristics and can therefore be generalised to other profiles. Dashed lines indicate the timing of vaccination. Shaded areas are 95% confidence intervals.

**Supplementary Figure 2g.**
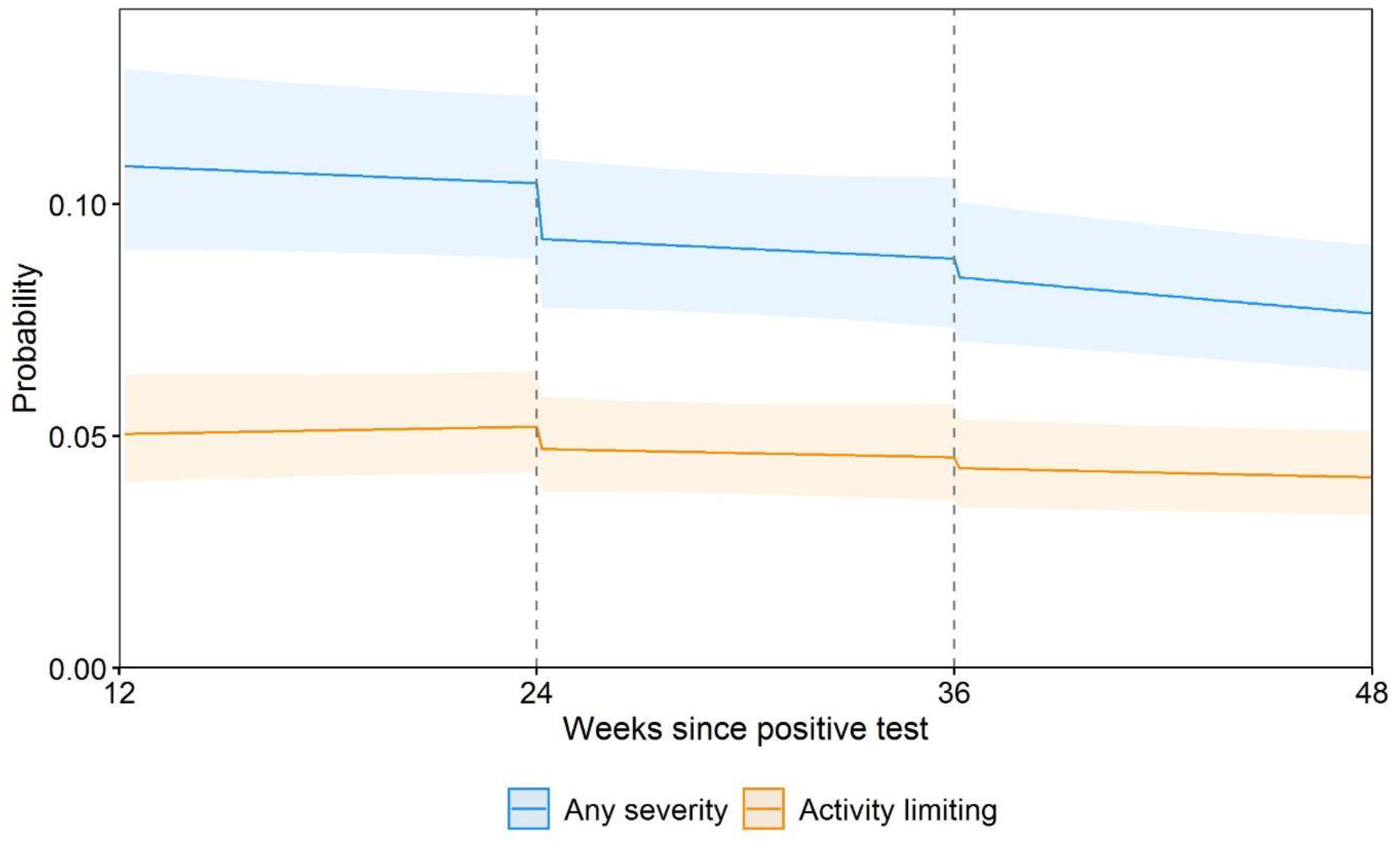
Modelled probabilities of Long Covid for a hypothetical study participant, sensitivity analysis 7: excluding participants infected in the first wave of the pandemic. Notes: This analysis was based on 19,085 participants (compared with 28,356 in the main analysis) infected from 11 September 2020 onwards. Probabilities are shown for a participant aged 50 years and in the modal group for other covariates (female, white, living in London, in an area in the least deprived quintile group, not a patient-facing health or social care worker, no pre-existing health conditions, not hospitalised at the acute phase of infection, and infected on 7 September 2020). While the estimated probabilities are specific to this profile, the proportional changes in probabilities after vaccination do not vary across characteristics and can therefore be generalised to other profiles. Dashed lines indicate the timing of vaccination. Shaded areas are 95% confidence intervals.

**Supplementary Figure 2h.**
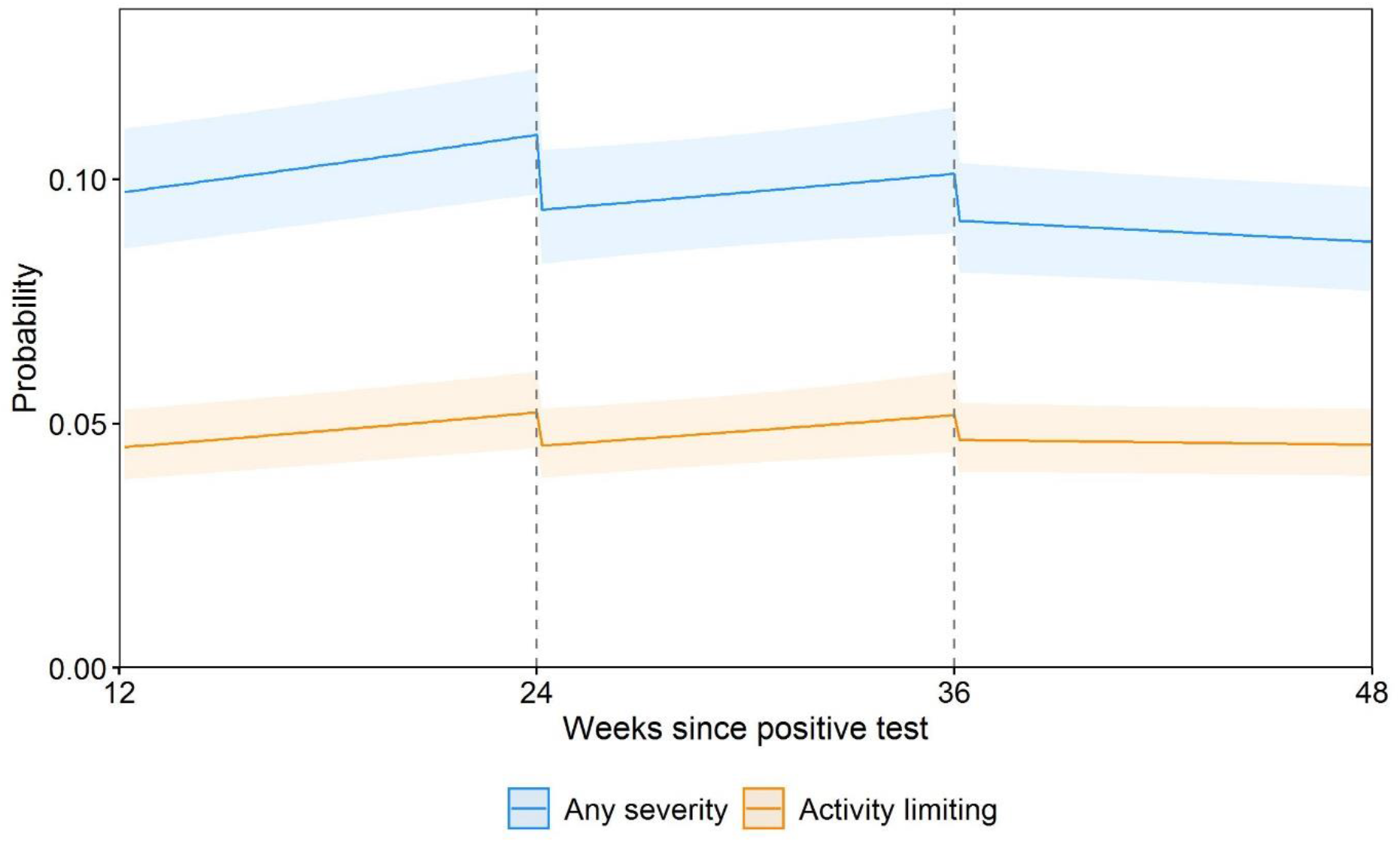
Modelled probabilities of Long Covid for a hypothetical study participant, sensitivity analysis 8: infection date for participants whose time of infection was determined by symptom onset >14 days before a positive swab reset to date of first positive swab. Notes: The infection date was moved forward by a median of 249 days among 698 participants (2.5%) whose time of infection was determined by symptom onset (rather than a confirmatory test) and was >14 days before a positive swab. Probabilities are shown for a participant aged 50 years and in the modal group for other covariates (female, white, living in London, in an area in the least deprived quintile group, not a patient-facing health or social care worker, no pre-existing health conditions, not hospitalised at the acute phase of infection, and infected on 7 September 2020). While the estimated probabilities are specific to this profile, the proportional changes in probabilities after vaccination do not vary across characteristics and can therefore be generalised to other profiles. Dashed lines indicate the timing of vaccination. Shaded areas are 95% confidence intervals.

**Supplementary Figure 3a.**
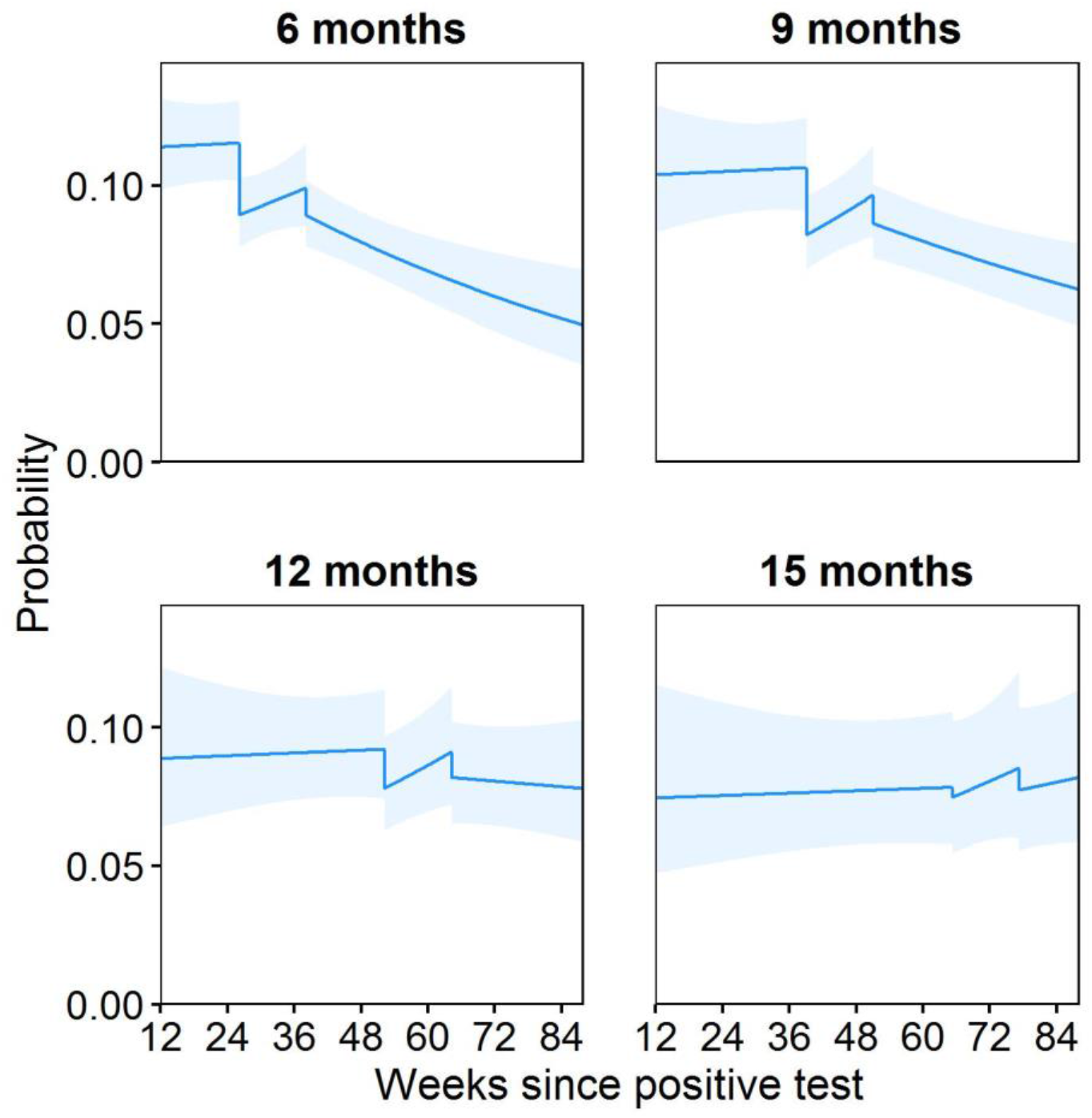
Modelled probabilities of Long Covid of any severity for hypothetical study participants who received their first vaccination 6, 9, 12, and 15 months after infection. Notes: Probabilities are shown for a participant of approximately mean age (50 years) and in the modal group for other covariates (female, white, living in London, in an area in the least deprived quintile group, not a patient-facing health or social care worker, no pre-existing health conditions, not hospitalised at the acute phase of infection, and infected on 7 September 2020). While the estimated probabilities are specific to this profile, the proportional changes in probabilities after vaccination do not vary across characteristics and can therefore be generalised to other profiles. Dashed lines indicate the timing of vaccination. Shaded areas are 95% confidence intervals. Probabilities were obtained by interacting all four exposure variables (changes in level and slope after each dose) with duration from infection to first vaccination (modelled as a restricted cubic spline).

**Supplementary Figure 3b.**
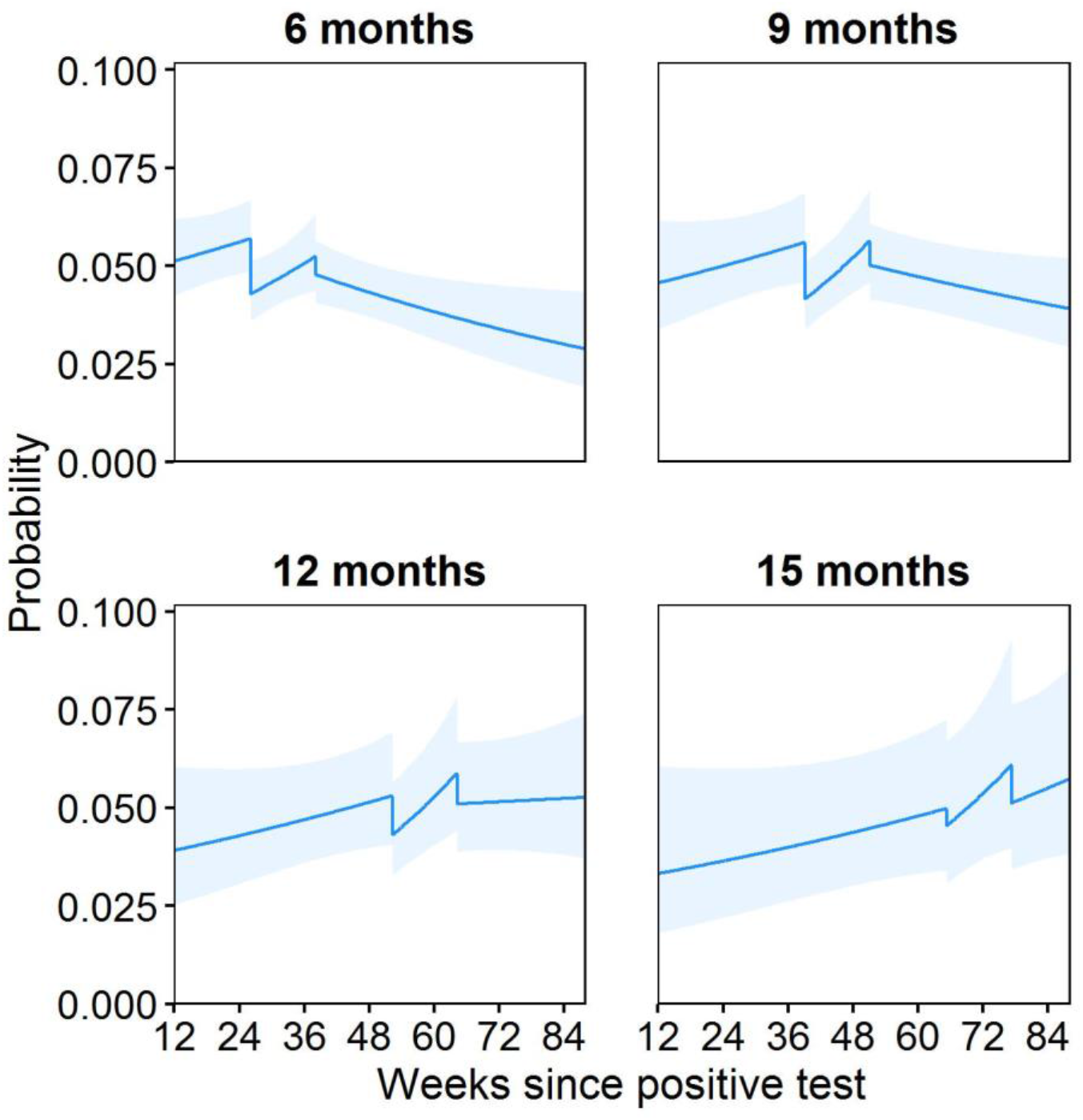
Modelled probabilities of activity-limiting Long Covid for hypothetical study participants who received their first vaccination 6, 9, 12, and 15 months after infection. Notes: Probabilities are shown for a participant of approximately mean age (50 years) and in the modal group for other covariates (female, white, living in London, in an area in the least deprived quintile group, not a patient-facing health or social care worker, no pre-existing health conditions, not hospitalised at the acute phase of infection, and infected on 7 September 2020). While the estimated probabilities are specific to this profile, the proportional changes in probabilities after vaccination do not vary across characteristics and can therefore be generalised to other profiles. Dashed lines indicate the timing of vaccination. Shaded areas are 95% confidence intervals. Probabilities were obtained by interacting all four exposure variables (changes in level and slope after each dose) with duration from infection to first vaccination (modelled as a restricted cubic spline).

