## Supplementary material for "Changes in the trajectory of Long Covid symptoms following COVID-19 vaccination: community-based cohort study": STROBE checklist

**Supplementary Appendix: Completed STROBE checklist**

| **Item** | **No.** | **Recommendation** | **Page** |
| --- | --- | --- | --- |
| **Title and abstract** | | | |
| Title and abstract | 1 | (*a*) Indicate the study's design with a commonly used term in the title or the abstract | 1 |
|  |  | (*b*) Provide in the abstract an informative and balanced summary of what was done and what was found | 2 |
| **Introduction** | | | |
| Background/rationale | 2 | Explain the scientific background and rationale for the investigation being reported | 4 |
| Objectives | 3 | State specific objectives, including any prespecified hypotheses | 4 |
| **Methods** | | | |
| Study design | 4 | Present key elements of study design early in the paper | 5 |
| Setting | 5 | Describe the setting, locations, and relevant dates, including periods of recruitment, exposure, follow-up, and data collection | 5 |
| Participants | 6 | Give the eligibility criteria, and the sources and methods of selection of participants | 5 |
| Variables | 7 | Clearly define all outcomes, exposures, predictors, potential confounders, and effect modifiers. Give diagnostic criteria, if applicable | 5-6 |
| Data sources/ measurement | 8 | For each variable of interest, give sources of data and details of methods of assessment (measurement). Describe comparability of assessment methods if there is more than one group | 4-5 |
| Bias | 9 | Describe any efforts to address potential sources of bias | 6 |
| Study size | 10 | Explain how the study size was arrived at | 5 |
| Quantitative variables | 11 | Explain how quantitative variables were handled in the analyses. If applicable, describe which groupings were chosen and why | 6, Suppl. Table 3 |
| Statistical methods | 12 | (*a*) Describe all statistical methods, including those used to control for confounding | 6 |
|  |  | (*b*) Describe any methods used to examine subgroups and interactions | 6 |
|  |  | (*c*) Explain how missing data were addressed | N/A |
|  |  | (*d*) If applicable, explain how loss to follow-up was addressed | N/A |
|  |  | (*e*) Describe any sensitivity analyses | 6-7 |
| **Results** | | | |
| Participants | 13 | (*a*) Report numbers of individuals at each stage of study, e.g. numbers potentially eligible, examined for eligibility, confirmed eligible, included in the study, completing follow-up, and analysed | 7 |
|  |  | (*b*) Give reasons for non-participation at each stage | 7 |
|  |  | (*c*) Consider use of a flow diagram | Suppl. Figure 1 |
| Descriptive data | 14 | (*a*) Give characteristics of study participants (e.g. demographic, clinical, social) and information on exposures and potential confounders | Table 1 |
|  |  | (*b*) Indicate number of participants with missing data for each variable of interest | N/A |
|  |  | (*c*) Summarise follow-up time (e.g. average and total amount) | 7 |
| Outcome data | 15 | Report numbers of outcome events or summary measures over time | 7 |
| Main results | 16 | (*a*) Report the numbers of individuals at each stage of the study, e.g. numbers potentially eligible, examined for eligibility, confirmed eligible, included in the study, completing follow-up, and analysed | 7 |
|  |  | (*b*) Give reasons for non-participation at each stage | 7 |
|  |  | (*c*) Consider use of a flow diagram | Suppl. Figure 1 |
| Other analyses | 17 | Report other analyses done, e.g. analyses of subgroups and interactions, and sensitivity analyses | 8 |
| **Discussion** | | | |
| Key results | 18 | Summarise key results with reference to study objectives | 8-9 |
| Limitations | 19 | Discuss limitations of the study, taking into account sources of potential bias or imprecision. Discuss both direction and magnitude of any potential bias | 10 |
| Interpretation | 20 | Give a cautious overall interpretation of results considering objectives, limitations, multiplicity of analyses, results from similar studies, and other relevant evidence | 10 |
| Generalisability | 21 | Discuss the generalisability (external validity) of the study results | 9-10 |
| **Other information** | | | |
| Funding | 22 | Give the source of funding and the role of the funders for the present study and, if applicable, for the original study on which the present article is based | 11 |
